# Cardiomyocyte-derived cell-free DNA in hypertrophic cardiomyopathy

**DOI:** 10.64898/2026.10.12.26365358

**Authors:** Daniel Kamna, Brett Davis, Cora E. Layman, Jack Wiedrick, Sammantha Avaylon, Kandace Wheeler, Israt Jahan, Lucia Carbone, Ahmad Masri

**Affiliations:** Knight Cardiovascular Institute, Oregon Health & Science University, Portland, OR, USA; Biostatistics and Design Program, Oregon Health & Science University, Portland, OR, USA; Department of Molecular and Medical Genetics, Oregon Health & Science University, Portland, OR, USA; Division of Genetics, Oregon National Primate Research Center, Beaverton, OR, USA

## Abstract

The hearts of patients with hypertrophic cardiomyopathy (HCM) are exposed to chronic wall stress and subclinical myocardial injury, yet conventional biomarkers such as cardiac troponin lack specificity for cardiomyocyte death. Here we evaluate whether cardiomyocyte-derived cell-free DNA (cfDNA-CM), quantified by whole-genome bisulfite sequencing and methylation-based cell-type deconvolution, can be measured in clinically stable HCM. We analyze 57 plasma samples from patients with HCM and 10 from patients with acute myocardial infarction as positive controls. Control samples show higher total cfDNA yield and cfDNA-CM fractions than HCM samples, supporting assay validity. In HCM, cfDNA-CM ranges from 0% to 6.4% of total cfDNA; higher cfDNA-CM is associated with higher body mass index, obesity and obstructive sleep apnea, but not with NT-proBNP or high-sensitivity troponin I. Quantifying cfDNA-CM in stable HCM is feasible, and its association with cardiometabolic burden independent of traditional biomarkers suggests that cfDNA-CM reflects a distinct domain of myocardial injury and remodeling.

## Introduction

Hypertrophic cardiomyopathy (HCM) affects approximately 1 in 200 individuals, making it the most common inherited cardiomyopathy^1^. This autosomal dominant disease, frequently caused by mutations of sarcomeric genes, most commonly myosin heavy chain and myosin binding protein C3, carries a substantial lifelong disease burden. Up to 62% of individuals diagnosed before age 40 develop atrial fibrillation, and nearly half experience symptomatic heart failure by age 60^2,3^.

Although HCM exhibits a heterogeneous clinical phenotype, its underlying pathophysiology is driven largely by pathological hypercontractility and impaired myocardial relaxation, leading to hypertrophy and adverse ventricular remodeling. These chronic changes lead to progressive wall stress and myocyte injury, which may be reflected by elevated levels of N-terminal pro-B-type natriuretic peptide (NT-proBNP) and cardiac troponin (cTn)^2^. cTn is considered the gold standard biomarker of myocyte injury and death, and represents an important diagnostic and prognostic tool^4^. However, it is chronically elevated in more than 50% of patients with HCM^5^.

The underlying pathophysiology that results in this persistent cTn elevation remains unclear, but may result from pressure overload, hypercontractility, or myocyte stretch rather than overt myocardial necrosis. Additionally, the complex biology of cTn is best illustrated by its increase in healthy individuals following intense physical activity, such as marathon running or prolonged cycling^6–8^.

Emerging data suggest that cTn release may occur through mechanisms beyond cardiomyocyte necrosis or apoptosis, including normal cell turnover, release of proteolytically cleaved troponin fragments, increased membrane permeability, and secretion via extracellular vesicles^9–11^. These complexities highlight the need for more specific biomarkers capable of distinguishing true cardiomyocyte injury from adaptive or non-cytotoxic processes in HCM.

Cell-free DNA (cfDNA), comprising extracellular DNA fragments circulating in bodily fluids such as plasma, urine, and cerebrospinal fluid, has emerged as a highly sensitive biomarker of cellular injury and death. Indeed, liquid biopsies using cfDNA have already established robust clinical utility in oncology detection, non-invasive prenatal testing, and solid-organ transplant rejection surveillance^12–14^. While total cfDNA levels fluctuate in cardiovascular conditions like hypertension and acute myocardial infarction (AMI), these conventional assays lack tissue specificity^15,16^. Because they measure total cfDNA, prior cardiovascular studies have been unable to differentiate the exact tissue of origin. Overcoming this limitation requires leveraging tissue-specific epigenetic marks, such as DNA methylation (the addition of a methyl group to cytosine residues, predominantly in CpG dinucleotides), to deconvolve and quantify the cardiomyocyte-specific cfDNA fraction.

Recent advances in next-generation sequencing have enabled genome-wide, high-resolution detection of DNA methylation across diverse sample types. The most widely adopted approach is whole-genome bisulfite sequencing (WGBS), in which unmethylated cytosines are converted to uracil (read as thymine) by bisulfite treatment, while methylated cytosines remain protected.

Because DNA methylation patterns are tissue-specific^17^, they can serve as molecular fingerprints to determine tissue of origin through comparison with established reference atlases^18^. Applied to cfDNA, WGBS therefore enables the assignment of circulating DNA fragments to their tissue of origin. This approach has been applied in diseases such as pulmonary hypertension and AMI^19,20^, but to the best of our knowledge, such an approach has not been employed to quantify cardiomyocyte-specific cfDNA in cardiomyopathies such as HCM.

In this study, we evaluated the feasibility of quantifying cardiomyocyte-specific cfDNA (cfDNA-CM) in patients with HCM and examined its association with traditional markers of disease severity. We hypothesized that circulating cfDNA-CM patterns would be distinct from those of conventional cardiac biomarkers (NT-proBNP and cTn) that are likely more reflective of wall stress and hypercontractility.

## Results

### Baseline characteristics of HCM and AMI patients

A total of 72 patients were enrolled in this study, including 62 patients with HCM and 10 with AMI (5 NSTEMI, 5 STEMI); 54 patients with HCM and all 10 patients with AMI were included in the final analysis. Three of the HCM patients contributed 2 samples each (57 HCM samples in total), separated in time by at least 1 year, and met stable enrollment criteria at both points of sampling. For the HCM cohort the mean age at time of sampling was 46.4 ± 16.6 years and 37 (64.9%) of the samples were from male patients. The median duration since HCM diagnosis was 5 years (IQR 2–14). A pathogenic variant was identified in 24 (42.1%) samples. A total of 42 (73.7%) samples were from patients who reported symptoms at the point of sampling, and 26 (45.6%) were associated with left ventricular outflow tract (LVOT) obstruction. Late gadolinium enhancement was present in 35 (61.4%) samples. The cohort had typical comorbidities seen in patients with HCM (Table 1). Median NT-proBNP was 396 pg/ml (IQR 169–825), with 35 (61.4%) having ≥300 pg/ml when sampled. Median troponin was 20 ng/L (IQR 7–118), and 32 (56.1%) had elevated values >10 ng/L when sampled. Baseline clinical, imaging, and biomarker characteristics are summarized in Table 1.

**Table 1.** Characteristics of HCM patient samples at the time of sampling, by tertile of total cfDNA yield.

| ***N (%)***<br>***mean*** ± ***SD***<br>***median (Q1, Q3) [min, max]*** | **Total**<br>*n* = 57 | **First Tertile**<br>*n* = 19 | **Second Tertile**<br>*n* = 19 | **Third Tertile**<br>*n* = 19 | ***P value*** |
|---|---|---|---|---|---|
| Male sex | 37 (64.9%) | 11 (57.9%) | 11 (57.9%) | 15 (78.9%) | 0.327 |
| Age at sample collection | 46.4 ± 16.6 | 44.4 ± 14.5 | 48.8 ± 17.8 | 45.8 ± 17.8 | 0.771 |
| Age > 50 years old | 23 (40.4%) | 4 (21.1%) | 10 (52.6%) | 9 (47.4%) | 0.119 |
| BMI (kg/m<sup>2</sup>) | 31.6 ± 5.5 | 32.3 ± 5.4 | 33.3 ± 5.6 | 29.1 ± 4.7 | 0.043 |
| Obese (BMI ≥ 30) | 33 (57.9%) | 11 (57.9%) | 16 (84.2%) | 6 (31.6%) | **0.006** |
| Diabetes mellitus | 2 (3.5%) | 0 (0.0%) | 2 (10.5%) | 0 (0.0%) | 0.321 |
| Coronary artery disease | 5 (8.8%) | 2 (10.5%) | 1 (5.3%) | 2 (10.5%) | >0.999 |
| Obstructive sleep apnea | 8 (14.0%) | 4 (21.1%) | 4 (21.1%) | 0 (0.0%) | 0.106 |
| Hypertension | 15 (26.3%) | 6 (31.6%) | 6 (31.6%) | 3 (15.8%) | 0.493 |
| Years since HCM diagnosis | 5 (2, 14)<br>[0, 34] | 5 (1, 14)<br>[0, 25] | 5 (0, 18)<br>[0, 34] | 5 (4, 12)<br>[0, 26] | 0.685 |
| NYHA class |  |  |  |  | 0.369 |
| *I* | 13 (22.8%) | 7 (36.8%) | 3 (15.8%) | 3 (15.8%) |  |
| *II* | 31 (54.4%) | 10 (52.6%) | 10 (52.6%) | 11 (57.9%) |  |
| *III/IV* | 13 (22.8%) | 2 (10.5%) | 6 (31.6%) | 5 (26.3%) |  |
| Family history of HCM | 27 (47.4%) | 8 (42.1%) | 11 (57.9%) | 8 (42.1%) | 0.668 |
| Pathological genetic mutation |  |  |  |  | 0.710 |
| *no* | 15 (26.3%) | 6 (31.6%) | 5 (26.3%) | 4 (21.1%) |  |
| *yes* | 24 (42.1%) | 8 (42.1%) | 6 (31.6%) | 10 (52.6%) |  |
| *unknown* | 18 (31.6%) | 5 (26.3%) | 8 (42.1%) | 5 (26.3%) |  |
| Late gadolinium enhancement |  |  |  |  | 0.332 |
| *no* | 12 (21.1%) | 2 (10.5%) | 5 (26.3%) | 5 (26.3%) |  |
| *yes* | 35 (61.4%) | 12 (63.2%) | 13 (68.4%) | 10 (52.6%) |  |
| *unknown* | 10 (17.5%) | 5 (26.3%) | 1 (5.3%) | 4 (21.1%) |  |
| Degree of fibrosis |  |  |  |  | 0.020 |
| *None* | 12 (25.5%) | 2 (14.3%) | 5 (27.8%) | 5 (33.3%) |  |
| *Mild* | 26 (55.3%) | 12 (85.7%) | 7 (38.9%) | 7 (46.7%) |  |
| *Moderate* | 7 (14.9%) | 0 (0.0%) | 6 (33.3%) | 1 (6.7%) |  |
| *Severe* | 2 (4.3%) | 0 (0.0%) | 0 (0.0%) | 2 (13.3%) |  |
| Symptomatic | 42 (73.7%) | 12 (63.2%) | 17 (89.5%) | 13 (68.4%) | 0.164 |
| Obstruction | 26 (45.6%) | 8 (42.1%) | 10 (52.6%) | 8 (42.1%) | 0.842 |
| AFIB | 5 (8.8%) | 1 (5.3%) | 3 (15.8%) | 1 (5.3%) | 0.602 |
| Implantable cardioverter-defibrillator | 25 (43.9%) | 7 (36.8%) | 6 (31.6%) | 12 (63.2%) | 0.127 |
| Myectomy | 13 (22.8%) | 3 (15.8%) | 2 (10.5%) | 8 (42.1%) | 0.086 |
| Hemoglobin (g/dL) (sex-adjusted) | 14.6 ± 1.0 | 14.7 ± 0.9 | 14.6 ± 1.0 | 14.4 ± 1.2 | 0.819 |
| Anemic (hemoglobin < 12[F],13.5[M]) | 5 (8.8%) | 1 (5.3%) | 1 (5.3%) | 3 (15.8%) | 0.602 |
| Creatinine (mg/dL) (sex-adjusted) | 0.9 ± 0.1 | 0.9 ± 0.2 | 0.9 ± 0.2 | 0.9 ± 0.1 | 0.738 |
| Elevated creatinine (>1.0[F],1.2[M]) | 7 (12.3%) | 3 (15.8%) | 1 (5.3%) | 3 (15.8%) | 0.678 |
| NT-proBNP (pg/mL) | 396 (169, 825)<br>[27, 4351] | 304 (58, 871)<br>[29, 1498] | 380 (213, 668)<br>[44, 3966] | 692 (260, 1769)<br>[27, 4351] | 0.145 |
| Elevated NT-proBNP (>300) | 35 (61.4%) | 10 (52.6%) | 12 (63.2%) | 13 (68.4%) | 0.702 |
| Troponin at collection (ng/L) | 20 (7, 118)<br>[3, 7775] | 20 (7, 92)<br>[3, 653] | 22 (8, 676)<br>[3, 7775] | 20 (7, 139)<br>[4, 3361] | 0.475 |
| Elevated troponin (>10) | 32 (56.1%) | 10 (52.6%) | 11 (57.9%) | 11 (57.9%) | >0.999 |

Clinical, demographic, imaging and laboratory characteristics of HCM samples (*n* = 57) stratified by tertile of total cfDNA yield. Data are *n* (%), mean ± SD or median (Q1, Q3) [min, max]. *P* values are from χ² or Fisher's exact tests (categorical variables) and Student's *t*-tests or Wilcoxon rank-sum tests (continuous variables); two-sided; *P* < 0.005 in bold. BMI, body mass index; HCM, hypertrophic cardiomyopathy; NYHA, New York Heart Association; AFIB, atrial fibrillation; NT-proBNP, N-terminal pro-B-type natriuretic peptide.

In comparison, the AMI cohort was older (70.8 ± 6.8 years), predominantly male (80.0%), and had a high prevalence of diabetes mellitus (70.0%), hypertension (70.0%), and obesity (70.0%). Mean left ventricular ejection fraction was 47.0 ± 12.5% in patients with NSTEMI and 41.0 ± 9.6% in patients with STEMI. Median troponin at collection was 5,926 ng/L (IQR 5,388–10,722) in NSTEMI and 49,200 ng/L (IQR 31,144–71,993) in STEMI. The AMI cohort served as a positive-control validation of the assay rather than a between-disease comparison. Baseline clinical and biomarker characteristics of both cohorts are summarized in Supplementary Table 1.

### cfDNA isolation and tissue deconvolution

cfDNA was isolated from all 67 plasma samples included in the final analysis and subsequently underwent WGBS and methylation-based cell-type deconvolution using a curated subset of 14 cell types from the 39-cell methylation atlas. Median total cfDNA yield was 12.22 ng (IQR 7.77– 18.94) in patients with HCM and 37.29 ng (IQR 22.96–64.80) in patients with AMI.

Across the study population, circulating cfDNA was predominantly derived from hematopoietic cells, with granulocytes and monocytes/macrophages representing the largest contributors.

Among samples from HCM patients, the predominant contributors were granulocytes (median 46.38%, IQR 38.85–55.47%), monocytes/macrophages (23.43%, IQR 17.09–30.05%), and endothelial cells (8.57%, IQR 4.59–12.71%). Median cardiomyocyte-derived and cardiac fibroblast-derived cfDNA fractions were 0.50% (IQR 0.00–1.52%) and 0.35% (IQR 0.00– 2.94%), respectively. Complete tissue deconvolution results are shown in Figs. 1 and 2.

**Fig. 1.**
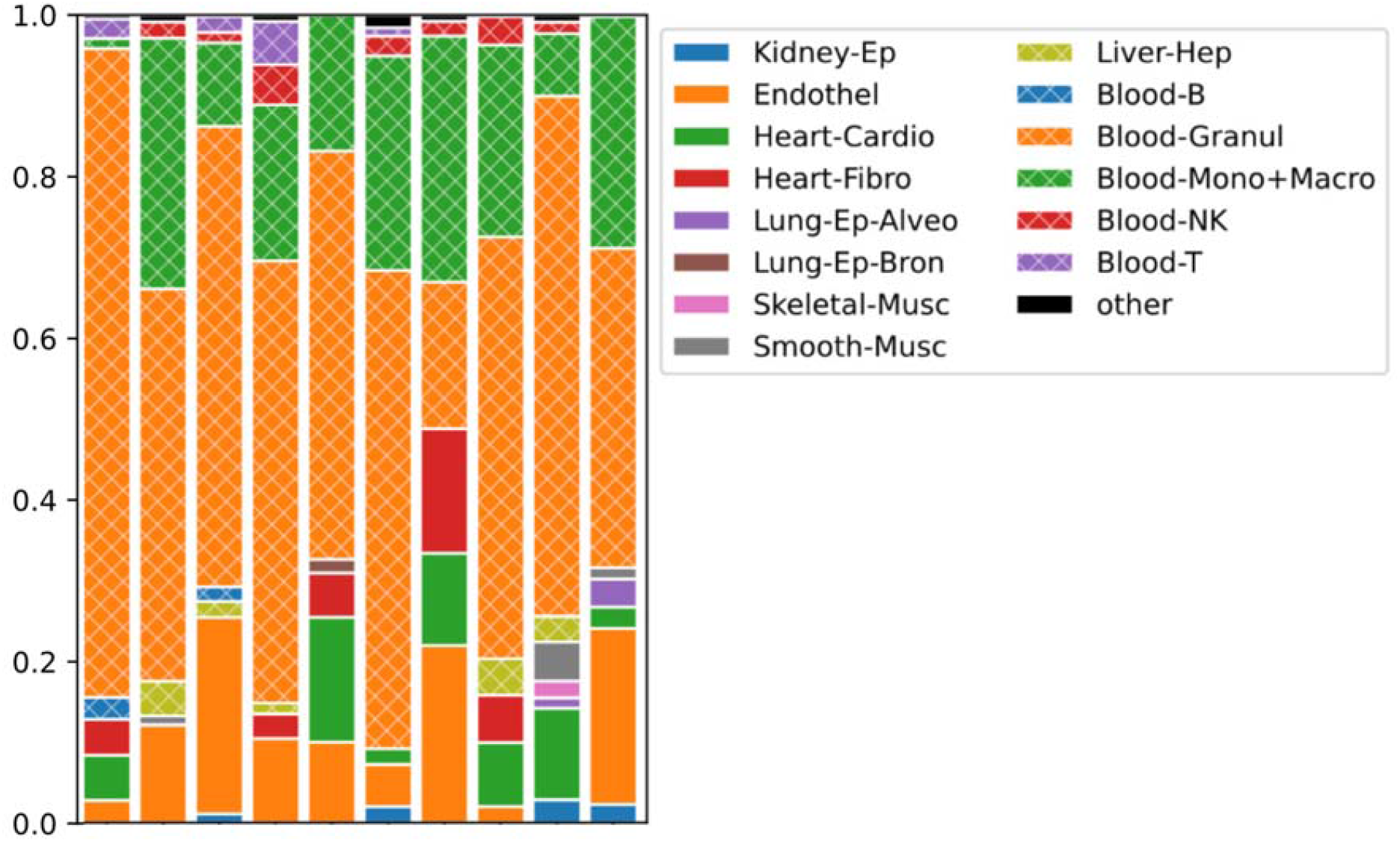
Methylation-based deconvolution of plasma cfDNA in patients with acute myocardial infarction. Stacked bars show estimated cell-type fractions of circulating cfDNA in individual acute myocardial infarction samples (*n* = 10 biologically independent samples; 5 NSTEMI and 5 STEMI). Colors indicate the cell-type categories shown in the legend. AMI, acute myocardial infarction; cfDNA, cell-free DNA; HCM, hypertrophic cardiomyopathy; NSTEMI, non-ST-segment-elevation myocardial infarction; STEMI, ST-segment-elevation myocardial infarction. Cell-type labels: Blood-B, B cells; Blood-Granul, granulocytes; Blood-Mono+Macro, monocytes/macrophages; Blood-NK, natural killer cells; Blood-T, T cells; Endothel, endothelial cells; Heart-Cardio, cardiomyocytes; Heart-Fibro, cardiac fibroblasts; Kidney-Ep, kidney epithelium; Liver-Hep, hepatocytes; Lung-Ep-Alveo, alveolar epithelium; Lung-Ep-Bron, bronchial epithelium; Skeletal-Musc, skeletal muscle; Smooth-Musc, smooth muscle

**Fig. 2.**
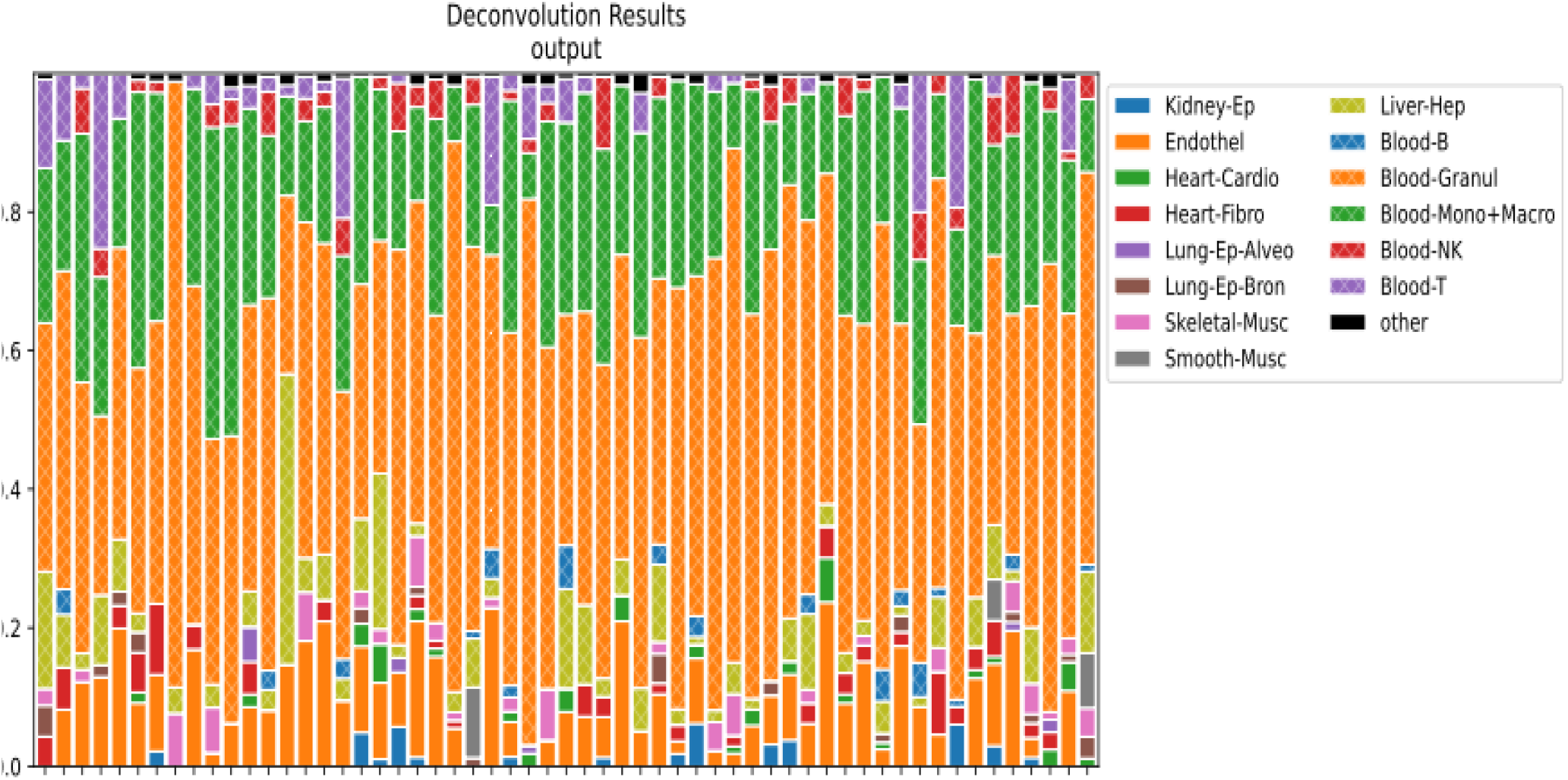
Methylation-based deconvolution of plasma cfDNA in patients with hypertrophic cardiomyopathy. Stacked bars show estimated cell-type fractions of circulating cfDNA in individual HCM samples (*n* = 57 samples from 54 patients). Colors indicate the cell-type categories shown in the legend. Cell-type labels: Blood-B, B cells; Blood-Granul, granulocytes; Blood-Mono+Macro, monocytes/macrophages; Blood-NK, natural killer cells; Blood-T, T cells; Endothel, endothelial cells; Heart-Cardio, cardiomyocytes; Heart-Fibro, cardiac fibroblasts; Kidney-Ep, kidney epithelium; Liver-Hep, hepatocytes; Lung-Ep-Alveo, alveolar epithelium; Lung-Ep-Bron, bronchial epithelium; Skeletal-Musc, skeletal muscle; Smooth-Musc, smooth muscle

### Biological and analytical validation of the methylation atlas

To assess whether the deconvolution approach detected the expected increase in cardiac-derived cfDNA during acute myocardial injury, cfDNA measures were compared across patients with HCM, NSTEMI, and STEMI (Table 2). Total cfDNA yield differed significantly across groups (*P* = 0.003), with median yields of 12.22 ng (IQR 7.77–18.94), 25.50 ng (IQR 22.96–64.80), and 39.48 ng (IQR 35.10–48.00) in HCM, NSTEMI, and STEMI, respectively. Cardiomyocyte-derived cfDNA fraction also differed significantly across groups (*P* = 0.002), with the highest fractions observed in patients with STEMI. Total heart-derived cfDNA fraction similarly differed across groups (*P* = 0.013), whereas cardiac fibroblast-derived cfDNA fraction did not (*P* = 0.775), underscoring that the group differences in total heart-derived cfDNA fraction were driven entirely by differing cardiomyocyte content. These findings supported the ability of the methylation-based deconvolution approach to detect cardiac-derived cfDNA in the setting of acute myocardial injury.

**Table 2.** cfDNA yield and cardiac-derived cfDNA percentages in HCM versus acute myocardial infarction samples.

| ***median (Q1, Q3) [min, max]*** | ***HCM (n=57)*** | ***NSTEMI (n=5)*** | ***STEMI (n=5)*** | ***P value*** |
|---|---|---|---|---|
| cfDNA yield (corrected), ng | 12.22 (7.77, 18.94) [5.33, 88.76] | 25.50 (22.96, 64.80) [9.97, 387.00] | 39.48 (35.10, 48.00) [14.56, 104.44] | 0.003 |
| Cardiomyocyte cfDNA percentage | 0.0 % (0.0, 2.0) [0.0, 6.0] | 0.0 % (0.0, 2.0) [0.0, 11.0] | 8.0 % (6.0, 11.0) [3.0, 15.0] | 0.002 |
| Cardiac fibroblast cfDNA percentage | 0.0 % (0.0, 3.0) [0.0, 10.0] | 0.0 % (0.0, 3.0) [0.0, 15.0] | 4.0 % (0.0, 5.0) [0.0, 6.0] | 0.775 |
| Total heart cfDNA percentage | 3.0 % (0.0, 4.0) [0.0, 11.0] | 2.0 % (0.0, 3.0) [0.0, 27.0] | 11.0 % (10.0, 14.0) [3.0, 21.0] | 0.013 |

Total cfDNA yield and estimated cardiomyocyte-, cardiac fibroblast- and total heart-derived cfDNA fractions in HCM (*n* = 57), NSTEMI (*n* = 5) and STEMI (*n* = 5) samples. Data are median (Q1, Q3) [min, max]. *P* values from Kruskal–Wallis/Wilcoxon rank-sum tests across groups; two-sided; *P* < 0.005 in bold. NSTEMI, non-ST-elevation myocardial infarction; STEMI, ST-elevation myocardial infarction.

In independent purified-tissue datasets, the methylation atlas correctly identified blood, heart, and liver as the predominant tissue sources in their respective reference samples, with mean recovered fractions of 94.3%, 68.2%, and 65.5%, respectively. In the in-silico mixture experiments, deconvolution consistently identified blood as the predominant tissue source.

Estimated heart and liver fractions showed variable agreement with the input proportions, and a portion of each mixture was assigned to the “other” category. Low-level cardiomyocyte signal was observed in nominally pure blood and liver samples, consistent with background classification noise and shared methylation features across tissues. These findings highlighted uncertainty in low-abundance cardiac cfDNA estimates and informed their interpretation in the clinical cohorts (Supplementary Figs. 1 and 2).

### Cross-method agreement

To determine whether cell-type proportion estimates were method-dependent, we compared UXM’s estimated cell type proportions with those from CelFiE-ISH across 50 HCM samples analyzed in a single batch with complete deconvolution results (Supplementary Table 2 and Supplementary Figs. 3 and 4). Agreement was highest for hepatocytes (CCC 0.87, Pearson’s r 0.88), with minimal bias (mean difference 0.009). Among the abundant blood cell types, agreement was highest for granulocytes (CCC 0.76, Pearson’s r 0.82) and T cells (CCC 0.71, Pearson’s r 0.79), followed by monocytes/macrophages (CCC 0.53, Pearson’s r 0.64). Mean differences for these three cell types were minimal (−0.015 to 0.009). Cardiomyocytes showed moderate concordance (CCC 0.50, Pearson’s r 0.50) with minimal bias (mean difference 0.002); however, limits of agreement (−3.4% to 3.8%) were wide relative to the mean estimated cardiomyocyte fractions (1.3–1.4%). Endothelial fraction estimates were consistently higher with CelFiE-ISH (mean difference 0.046; CCC 0.36, Pearson’s r 0.53), reflecting a systematic offset for this cell type with limits of agreement ranging from −7.7% to 17%. Agreement was poor for low abundance cell types (B cells, NK cells, alveolar and bronchial lung, cardiac fibroblasts, skeletal muscle, and smooth muscle) where all had CCC less than 0.15 and estimated fractions were close to zero in most samples. For skeletal muscle, smooth muscle, and cardiac fibroblasts, UXM generally estimated higher fractions than CelFiE-ISH. These findings indicate that while both methods produce broadly concordant estimates for abundant cell types, estimates for lower-abundance populations are method-dependent. UXM was used for the primary analyses because its performance had been independently evaluated using pure-tissue reference samples and simulated tissue mixtures.

### Total cfDNA yield and clinical associations

Patient samples were stratified into tertiles of total cfDNA yield (n = 19 per group). Despite substantial differences in median total cfDNA yield (6.59 ng vs 12.22 ng vs 24.15 ng; *P* < 0.001), cardiac-specific cfDNA fractions remained uniformly low and did not differ significantly across tertiles for any of the cardiomyocyte-derived, cardiac fibroblast-derived, or total heart-derived cfDNA fractions (all *P* > 0.1; Table 3, top block).

**Table 3.**
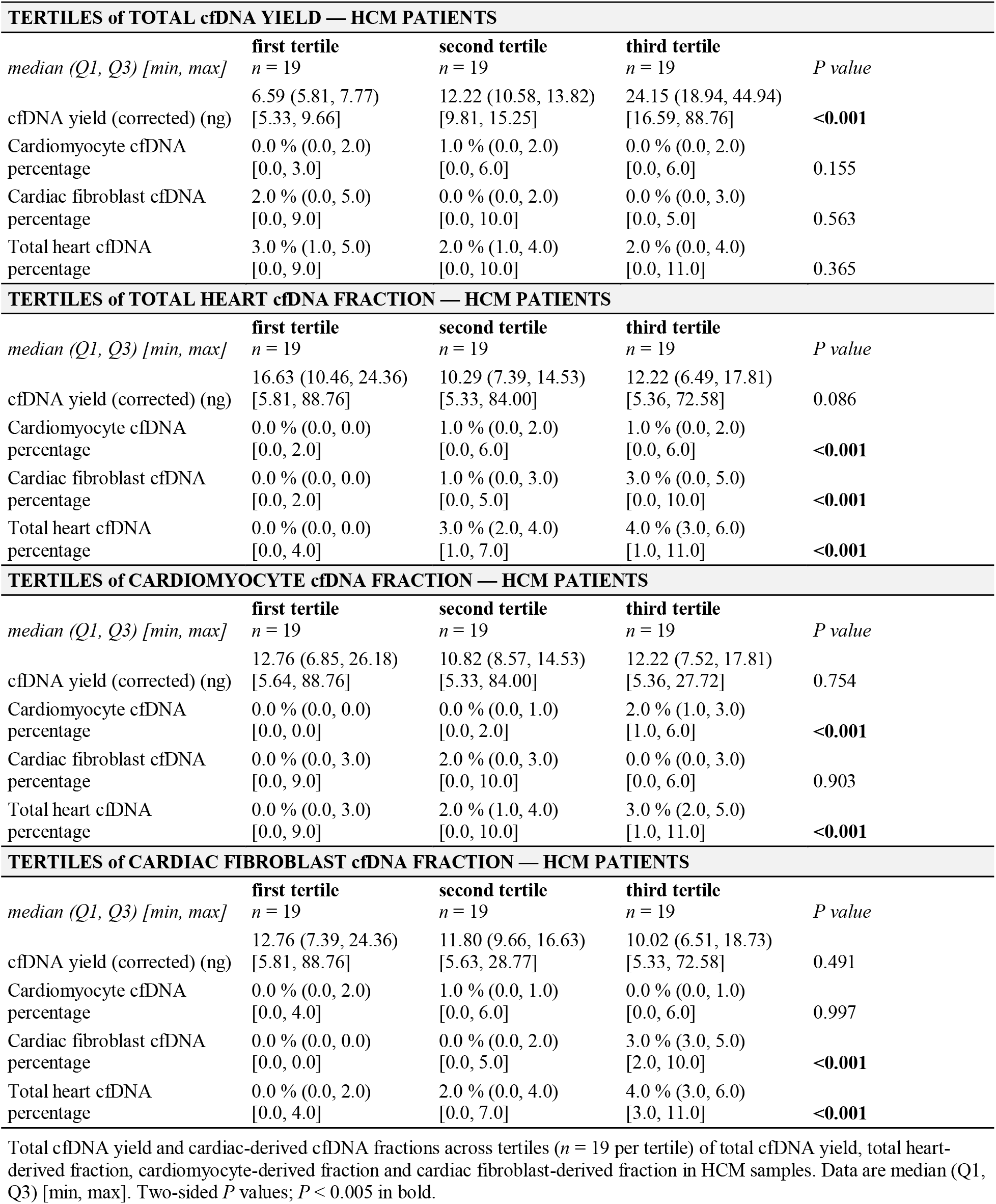
cfDNA measurements stratified by tertiles of total cfDNA yield and of cardiac cell-type percentages.

| **TERTILES of TOTAL cfDNA YIELD — HCM PATIENTS** |  |  |  |  |
| *median (Q1, Q3) [min, max]* | **first tertile**<br>*n* = 19 | **second tertile**<br>*n* = 19 | **third tertile**<br>*n* = 19 | *P value* |
|---|---|---|---|---|
| cfDNA yield (corrected) (ng) | 6.59 (5.81, 7.77)<br>[5.33, 9.66] | 12.22 (10.58, 13.82)<br>[9.81, 15.25] | 24.15 (18.94, 44.94)<br>[16.59, 88.76] | **<0.001** |
| Cardiomyocyte cfDNA percentage | 0.0 % (0.0, 2.0)<br>[0.0, 3.0] | 1.0 % (0.0, 2.0)<br>[0.0, 6.0] | 0.0 % (0.0, 2.0)<br>[0.0, 6.0] | 0.155 |
| Cardiac fibroblast cfDNA percentage | 2.0 % (0.0, 5.0)<br>[0.0, 9.0] | 0.0 % (0.0, 2.0)<br>[0.0, 10.0] | 0.0 % (0.0, 3.0)<br>[0.0, 5.0] | 0.563 |
| Total heart cfDNA percentage | 3.0 % (1.0, 5.0)<br>[0.0, 9.0] | 2.0 % (1.0, 4.0)<br>[0.0, 10.0] | 2.0 % (0.0, 4.0)<br>[0.0, 11.0] | 0.365 |
| **TERTILES of TOTAL HEART cfDNA FRACTION — HCM PATIENTS** |  |  |  |  |
| *median (Q1, Q3) [min, max]* | **first tertile**<br>*n* = 19 | **second tertile**<br>*n* = 19 | **third tertile**<br>*n* = 19 | *P value* |
| cfDNA yield (corrected) (ng) | 16.63 (10.46, 24.36)<br>[5.81, 88.76] | 10.29 (7.39, 14.53)<br>[5.33, 84.00] | 12.22 (6.49, 17.81)<br>[5.36, 72.58] | 0.086 |
| Cardiomyocyte cfDNA percentage | 0.0 % (0.0, 0.0)<br>[0.0, 2.0] | 1.0 % (0.0, 2.0)<br>[0.0, 6.0] | 1.0 % (0.0, 2.0)<br>[0.0, 6.0] | **<0.001** |
| Cardiac fibroblast cfDNA percentage | 0.0 % (0.0, 0.0)<br>[0.0, 2.0] | 1.0 % (0.0, 3.0)<br>[0.0, 5.0] | 3.0 % (0.0, 5.0)<br>[0.0, 10.0] | **<0.001** |
| Total heart cfDNA percentage | 0.0 % (0.0, 0.0)<br>[0.0, 4.0] | 3.0 % (2.0, 4.0)<br>[1.0, 7.0] | 4.0 % (3.0, 6.0)<br>[1.0, 11.0] | **<0.001** |
| **TERTILES of CARDIOMYOCYTE cfDNA FRACTION — HCM PATIENTS** |  |  |  |  |
| *median (Q1, Q3) [min, max]* | **first tertile**<br>*n* = 19 | **second tertile**<br>*n* = 19 | **third tertile**<br>*n* = 19 | *P value* |
| cfDNA yield (corrected) (ng) | 12.76 (6.85, 26.18)<br>[5.64, 88.76] | 10.82 (8.57, 14.53)<br>[5.33, 84.00] | 12.22 (7.52, 17.81)<br>[5.36, 27.72] | 0.754 |
| Cardiomyocyte cfDNA percentage | 0.0 % (0.0, 0.0)<br>[0.0, 0.0] | 0.0 % (0.0, 1.0)<br>[0.0, 2.0] | 2.0 % (1.0, 3.0)<br>[1.0, 6.0] | **<0.001** |
| Cardiac fibroblast cfDNA percentage | 0.0 % (0.0, 3.0)<br>[0.0, 9.0] | 2.0 % (0.0, 3.0)<br>[0.0, 10.0] | 0.0 % (0.0, 3.0)<br>[0.0, 6.0] | 0.903 |
| Total heart cfDNA percentage | 0.0 % (0.0, 3.0)<br>[0.0, 9.0] | 2.0 % (1.0, 4.0)<br>[0.0, 10.0] | 3.0 % (2.0, 5.0)<br>[1.0, 11.0] | **<0.001** |
| **TERTILES of CARDIAC FIBROBLAST cfDNA FRACTION — HCM PATIENTS** |  |  |  |  |
| *median (Q1, Q3) [min, max]* | **first tertile**<br>*n* = 19 | **second tertile**<br>*n* = 19 | **third tertile**<br>*n* = 19 | *P value* |
| cfDNA yield (corrected) (ng) | 12.76 (7.39, 24.36)<br>[5.81, 88.76] | 11.80 (9.66, 16.63)<br>[5.63, 28.77] | 10.02 (6.51, 18.73)<br>[5.33, 72.58] | 0.491 |
| Cardiomyocyte cfDNA percentage | 0.0 % (0.0, 2.0)<br>[0.0, 4.0] | 1.0 % (0.0, 1.0)<br>[0.0, 6.0] | 0.0 % (0.0, 1.0)<br>[0.0, 6.0] | 0.997 |
| Cardiac fibroblast cfDNA percentage | 0.0 % (0.0, 0.0)<br>[0.0, 0.0] | 0.0 % (0.0, 2.0)<br>[0.0, 5.0] | 3.0 % (3.0, 5.0)<br>[2.0, 10.0] | **<0.001** |
| Total heart cfDNA percentage | 0.0 % (0.0, 2.0)<br>[0.0, 4.0] | 2.0 % (0.0, 4.0)<br>[0.0, 7.0] | 4.0 % (3.0, 6.0)<br>[3.0, 11.0] | **<0.001** |

Total cfDNA yield and cardiac-derived cfDNA fractions across tertiles (*n* = 19 per tertile) of total cfDNA yield, total heart-derived fraction, cardiomyocyte-derived fraction and cardiac fibroblast-derived fraction in HCM samples. Data are median (Q1, Q3) [min, max]. Two-sided *P* values; *P* < 0.005 in bold.

Overall, clinical characteristics were largely similar across tertiles (Table 1). The total cfDNA yield was not associated with established markers of disease severity or myocardial injury.

Specifically, NT-proBNP levels did not differ significantly across the lowest, middle, and highest cfDNA tertiles, with median concentrations of 304 pg/mL (IQR 58–871), 380 pg/mL (IQR 213–668), and 692 pg/mL (IQR 260–1769), respectively (*P* = 0.145). The proportion of patients with elevated NT-proBNP (>300 pg/mL) was likewise comparable across tertiles, representing 52.6% of the first tertile, 63.2% of the second tertile, and 68.4% of the third tertile (*P* = 0.702). Similarly, median troponin concentrations at collection were similar across the three tertiles at 20 ng/L (IQR 7–92), 22 ng/L (IQR 8–676), and 20 ng/L (IQR 7–139) (*P* = 0.475), with no significant difference in the prevalence of elevated troponin (>10 ng/L) between the groups (52.6% vs 57.9% vs 57.9%, respectively; *P* > 0.999).

Among baseline characteristics, BMI differed significantly across tertiles (32.3 ± 5.4 kg/m^2^ vs 33.3 ± 5.6 kg/m^2^ vs 29.1 ± 4.7 kg/m^2^; *P* = 0.043), with corresponding differences in obesity prevalence (57.9% vs 84.2% vs 31.6%; *P* = 0.006). Furthermore, myocardial fibrosis severity on cardiac magnetic resonance (CMR) differed significantly across tertiles (*P* = 0.020). Moderate fibrosis was most prevalent in the middle tertile (33.3%), whereas severe fibrosis was seen only in the highest tertile (13.3%).

### Myocardial-derived cfDNA fraction and clinical associations

Patients contributing samples in the highest total myocardial cfDNA tertile (n = 19) were younger (mean age 38.3 ± 14.4 years) at the point of sampling compared with those in the lowest tertile (50.5 ± 19.5 years; *P* = 0.042). BMI increased across tertiles (28.4 ± 4.6 vs 32.3 ± 4.9 vs 34.0 ± 5.6 kg/m²; *P* = 0.004), with a significantly higher prevalence of obesity in the upper tertiles (26.3% vs 73.7% vs 73.7%; *P* = 0.004) (Supplementary Table 4).

Structural markers of disease also differed across tertiles. Late gadolinium enhancement differed across tertiles (*P* = 0.039), and the prevalence of LVOT obstruction was significantly higher in the middle and highest tertiles (*P* = 0.031). Fibrosis severity did not differ significantly across groups (*P* = 0.270). There were no significant differences in other characteristics, including NT-proBNP (*P* = 0.126) and troponin (*P* = 0.485) (Supplementary Table 4).

Similar findings were seen when cardiomyocyte cfDNA fraction was investigated instead of total myocardial cfDNA. Higher cardiomyocyte cfDNA fraction was associated with higher BMI (28.2 ± 5.0 vs 32.2 ± 4.5 vs 34.2 ± 5.4 kg/m²; *P* = 0.001) and with obesity prevalence (21.1% vs 73.7% vs 78.9%; *P* < 0.001). Obstructive sleep apnea was also more common in the highest tertile (31.6%) compared with the lower tertiles (5.3% each; *P* = 0.038). These associations suggest a possible relationship between cardiometabolic burden and cardiomyocyte-derived cfDNA release. There were non-significant trends towards more symptoms, LVOT obstruction, atrial fibrillation and late gadolinium enhancement in samples with higher cardiomyocyte cfDNA (Supplementary Table 5).

Cardiomyocyte-derived cfDNA fraction was not associated with NT-proBNP (*P* = 0.827), median troponin concentration (*P* = 0.293), or elevated troponin status (*P* = 0.127).

Per-sample relationships are shown in Fig. 3. Cardiomyocyte-derived cfDNA fraction was higher in STEMI than in HCM or NSTEMI samples (Kruskal–Wallis *P* = 0.002; Fig. 3a). Within HCM (*n* = 57), cardiomyocyte-derived cfDNA fraction correlated positively with BMI (Spearman ρ = 0.42, *P* = 0.001; Fig. 3b) and was higher in samples from patients with obesity than in those without (median 1.02% vs 0.00%; *P* < 0.001; Fig. 3c), whereas cardiac fibroblast-derived cfDNA fraction did not correlate with BMI (ρ = 0.01, *P* = 0.93). By contrast, cardiomyocyte-derived cfDNA fraction showed no correlation with hs-cTnI (ρ = −0.13, *P* = 0.34; Fig. 3d) or NT-proBNP (ρ = −0.08, *P* = 0.55; Fig. 3e).

**Fig. 3.**
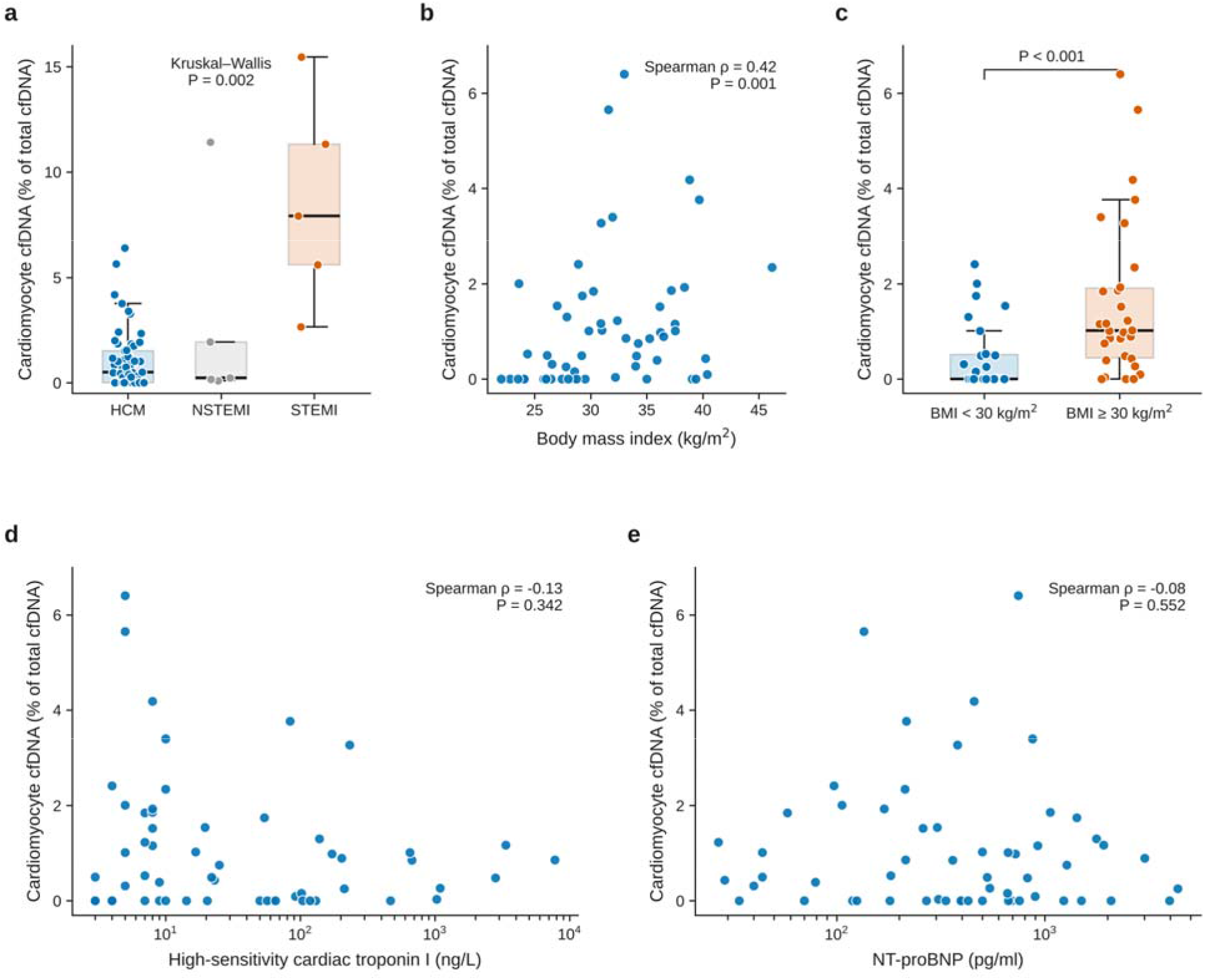
Cardiomyocyte-derived cfDNA is elevated in acute myocardial infarction and, in HCM, is associated with body mass index but not with conventional cardiac biomarkers. **a**, Cardiomyocyte-derived cfDNA fraction (% of total cfDNA) in HCM **b**, Cardiomyocyte-derived cfDNA fraction versus body mass index (BMI) in HCM samples **c**, Cardiomyocyte-derived cfDNA fraction in HCM samples from patients without and with obesity (BMI ≥ 30 kg/m^2^) **d**,**e**, Cardiomyocyte-derived cfDNA fraction versus high-sensitivity cardiac troponin I (**d**) and NT-proBNP (**e**) in HCM samples, both on logarithmic axes. BMI, body mass index; cfDNA, cell-free DNA; HCM, hypertrophic cardiomyopathy; hs-cTnI, high-sensitivity cardiac troponin I; NSTEMI, non-ST-segment-elevation myocardial infarction; NT-proBNP, N-terminal pro-B-type natriuretic peptide; STEMI, ST-segment-elevation myocardial infarction; ρ, Spearman rank correlation coefficient.

Finally, cardiac fibroblast-derived cfDNA fraction was not significantly associated with BMI, obesity, or most cardiometabolic comorbidities (all *P* > 0.05; Supplementary Table 6).

## Discussion

In this study, we systematically characterized circulating cfDNA in a well-phenotyped cohort of patients with HCM, demonstrating the feasibility of quantifying different sources of cfDNA using methylation-based deconvolution. Three principal findings emerged. First, quantification of cfDNA-CM in patients with HCM who are not suffering from an acute illness is feasible.

Second, higher cfDNA-CM levels were associated with increased BMI and obesity, potentially linking cardiometabolic burden to circulating markers of myocardial cell turnover. Third, cfDNA-CM fractions were not associated with established biomarkers of myocardial wall stress and cellular injury, including NT-proBNP and hs-cTnI. These findings suggest that cfDNA-CM may represent a distinct biological domain not captured by traditional circulating biomarkers.

Cell-free DNA has emerged as a powerful biomarker of tissue injury and cellular turnover. Released into blood circulation following cell death, cfDNA provides a dynamic snapshot of ongoing biological processes occurring throughout the body. Unlike conventional protein biomarkers, which might be released through multiple pathways, cfDNA directly originates from cells undergoing injury or turnover^29–31^. Additionally, a unique characteristic of cfDNA is that it retains tissue-specific methylation patterns that permit identification of its cellular origin^32^. Using the tissue-specific characteristics of cfDNA to assess disease state has already demonstrated clinical utility in fields such as oncology, prenatal diagnostics, and transplant medicine, and has shown promise in acute cardiovascular conditions including myocardial infarction^12–14,19^. By enabling direct quantification of cardiomyocyte-derived DNA fragments in the circulation, methylation-based cfDNA analysis offers a unique opportunity to assess myocardial injury and remodeling through a biologically distinct lens that may complement traditional biomarkers.

The association between cfDNA-CM and BMI/obesity is notable because, to our knowledge, it has not previously been described and, importantly, was not observed with traditional biomarkers. While this novel observation requires confirmation in a larger independent cohort, it can be leveraged to guide future studies to determine the possible mechanism behind the association between cfDNA-CM and BMI/obesity. Obesity is a well-established driver of systemic inflammation and increased hemodynamic load, both of which may exacerbate the underlying pathophysiology of HCM. As described by Spudich, HCM mutations that drive the hypercontractility in HCM act through altered ATPase kinetics and the recruitment of excess myosin heads into an active state, creating a baseline of high energetic demand^33^. Prior work by Fumagalli et al. has demonstrated that obesity in HCM is independently associated with increased left ventricular wall stress and adverse structural remodeling, which we believe superimposes an additional hemodynamic load onto an already inefficient system^34^. We speculate that the observed increase in cfDNA-CM may represent the molecular ‘breaking point’ of a heart pushed to its metabolic capacity. This ‘multi-hit’ stressor likely amplifies metabolic demand, promoting oxidative stress and the generation of reactive oxygen species (ROS). Recent evidence from Zhuo et al. further links these oxidative stress-related transcriptional programs in HCM to apoptotic signaling pathways^35^. We therefore suggest that chronic metabolic and hemodynamic stress promotes a low-grade, ROS-mediated cardiomyocyte injury, resulting in an accelerated but ‘silent’ cellular turnover. In this context, cfDNA-CM emerges as a sensitive, real-time genomic indicator of metabolic exhaustion and subclinical cellular attrition, a signal that traditional protein biomarkers of overt necrosis may overlook. Consistent with this cardiometabolic phenotype, cfDNA-CM was also associated with obstructive sleep apnea. These findings raise the intriguing possibility that cardiometabolic burden may contribute to subclinical cardiomyocyte turnover or DNA release, independent of overt myocardial injury detectable by conventional biomarkers. By contrast, cardiac fibroblast-derived cfDNA (f-cfDNA) did not show similar associations, suggesting that fibroblast DNA release may be more tightly linked to structural remodeling or extracellular matrix turnover than metabolic stress per se.

An equally intriguing observation is the lack of correlation between cfDNA-CM and established markers of injury like hs-cTnI and NT-proBNP. While NT-proBNP is a sensitive indicator of ventricular stretch and remodeling in HCM, it does not necessarily reflect the rate of myocardial cell death. In our cohort, the lack of correlation between NT-proBNP and cfDNA-CM suggests that a patient can have significant hemodynamic stress (high stretch) without necessarily crossing the threshold into the accelerated, ROS-mediated cellular turnover (high cfDNA-CM) that we observed in the setting of obesity. The lack of correlation between cfDNA-CM and troponin, even in a cohort specifically enriched for chronically elevated troponin, highlights a potentially important ‘biomarker uncoupling’ in HCM. While these markers typically increase in tandem during acute myocardial injury^19^, our data suggest that in HCM, they reflect distinct cellular release pathways.

One possibility for this discrepancy is the differential partitioning of genomic and protein cargo into extracellular vesicles (EVs). While the presence of circulating cfDNA-CM typically implies lack of cellular integrity or true cell death, the associated troponin may be sequestered rather than free-floating. Recent evidence indicates that apoptosis triggers a substantial release of large EVs (LEVs), which carry a distinct proteomic and genomic signature^36^. This is supported by the work of Spanos et al., which demonstrated that in HCM, a substantial fraction of circulating troponin is sequestered within EVs, whereas troponin in acute syndromes remains primarily free-floating^37^. Because conventional high-sensitivity assays primarily detect non-vesicle-associated protein, the EV-bound troponin remains effectively ‘masked’ from standard detection.

Furthermore, while Spanos et al. suggest that EV-associated troponin may represent an active secretory response to myocardial stress, the surge in cfDNA-CM we observed likely reflects a high-volume wave of cellular attrition where genomic material is released alongside these apoptotic vesicles. Consequently, cfDNA-CM may serve as a more direct indicator of actual cardiomyocyte death, capturing a ‘silent’ turnover that is biologically and analytically distinct from the active secretory pathways that drive chronic troponin elevation in HCM.

The lack of correlation between cfDNA-CM and hs-cTnI in our HCM cohort may further be influenced by the presence of autoantibodies to cardiac troponin I. Anti-cardiac troponin antibodies have been found in up to 20% of healthy individuals and up to 27.6% in those with cardiomyopathies^38,39^. As demonstrated by Eriksson et al., autoantibodies can effectively mask the release of troponin from injured tissue, particularly in assays that target the mid-fragment of the protein. At high titers, these antibodies can block the signal of small amounts of circulating troponin, leading to a significant delay or total failure in detection^40^. Conversely, anti-cardiac troponin autoantibodies may also form circulating macrotroponin immune complexes that increase molecular size and prolong plasma half-life, potentially leading to persistently elevated hs-cTnI levels despite relatively low ongoing cardiomyocyte injury or turnover^41^. Although anti-troponin antibodies were not measured in our cohort and their contribution cannot be determined, the hs-cTnI concentrations analyzed in this study represent the clinically measured values used in patient assessment, risk stratification, and increasingly as biomarkers of treatment response in HCM clinical trials. Thus, regardless of the mechanisms influencing circulating troponin concentrations, the observed dissociation between clinically measured hs-cTnI and cfDNA-CM supports the possibility that cfDNA-CM provides complementary biological information not captured by conventional troponin measurement. Whereas hs-cTnI is classically interpreted as a marker of acute or chronic cardiomyocyte injury, cfDNA-CM may instead capture ongoing cellular turnover or remodeling. Together, these findings challenge the assumption that traditional cardiac biomarkers and cfDNA reflect the same underlying pathophysiology and support a complementary role for cfDNA as a marker of myocardial stress.

The lack of association between cardiac fibroblast-derived cfDNA and LGE or fibrosis severity raises the possibility that circulating fibroblast-derived DNA reflects active fibroblast turnover or activation rather than the cumulative burden of replacement fibrosis captured by LGE. Future longitudinal studies integrating f-cfDNA with markers of fibroblast activation may help determine whether it provides a dynamic measure of active myocardial remodeling.

From a translational perspective, our findings have several implications. First, cfDNA profiling could provide a noninvasive window into myocardial biology, offering opportunities for early detection of cellular stress before the development of irreversible hypertrophy or fibrosis.

Second, the disconnect between cfDNA and traditional biomarkers suggests that current clinical metrics may underestimate subclinical myocardial injury or remodeling in certain patient subsets, particularly those with metabolic comorbidities. Third, these results lay the groundwork for integrating cfDNA analyses with genetic and imaging studies to better stratify risk and monitor disease progression in genotype-positive individuals. Additionally, the emergence of novel disease-modifying therapies highlights the clinical utility of differentiating physiologic biomarker shifts from true cell death. For example, the cardiac myosin activator omecamtiv mecarbil demonstrated favorable outcomes in the GALACTIC-HF trial but faced regulatory hurdles due to concerns regarding drug-related elevations in troponin^42^. Although this troponin rise originally raised alarms for cardiotoxicity, post-hoc analyses suggested it did not reflect myocardial necrosis^43^. In such scenarios, cfDNA could serve as a useful adjunctive biomarker, offering a more specific readout of actual cell death to clarify ambiguous troponin elevations and de-risk promising therapeutics. A similar challenge has emerged with gene therapy in cardiomyopathies and neuromuscular disorders characterized by chronically elevated troponin.

In the EMBARK trial of delandistrogene moxeparvovec for Duchenne muscular dystrophy, myocarditis occurred as a treatment-related serious adverse event following gene therapy^44^. In this setting, where baseline troponin may already be elevated and skeletal muscle involvement may further complicate interpretation of circulating biomarkers, identifying treatment-emergent myocardial injury can be particularly challenging. Cardiomyocyte-derived cfDNA could provide an opportunity to distinguish changes in baseline biomarker profiles from true cardiomyocyte injury, potentially facilitating the evaluation and monitoring of myocarditis or other cardiac toxicities during gene therapy. More broadly, this approach may advance our understanding of myocardial turnover in vivo and establish cfDNA as a clinically useful biomarker across a range of chronic cardiomyopathies, extending beyond HCM to other conditions characterized by progressive remodeling and heart failure.

This study has limitations. It was a single-center pilot study designed to assess the feasibility of quantifying cfDNA with its cellular origin in clinically stable outpatients with HCM. The sampling design was not fully independent, with a few examples of multiple samples contributed by the same patients. This was done to increase the sample size overall in view of our sampling time constraints for this pilot study. There were only a small number of repeated samples (3 of 54 patients contributed two samples each; the remaining 51 contributed one), and we took great care to make sure the repeated samples were separated by at least 1 year (which in our HCM cohort often translated to at least some changes in clinical presentation), but this poses a risk of mildly inflated associations owing to double-counting. Because of the limitations imposed by our small sample size, larger and better-powered studies are needed to confirm our findings and establish more robust correlations with underlying disease pathology. We used disease controls rather than healthy volunteers because little myocardial cfDNA is expected in healthy individuals. Additionally, the cross-sectional design precludes inference regarding temporal dynamics of cfDNA release relative to disease progression. Anti-cardiac troponin antibodies and extracellular vesicle-associated troponin were not measured, limiting our ability to determine whether these factors contributed to the observed dissociation between hs-cTnI and cfDNA-CM. The association between cfDNA-CM and BMI was sensitive to the deconvolution algorithm and marker depth, and therefore requires confirmation in an independent cohort. Future studies incorporating longitudinal sampling, broader genotypic diversity, and mechanistic interrogation of cfDNA sources are needed to clarify the biological and clinical relevance of our observations. In summary, estimation of cardiac-specific cfDNA in HCM is feasible and may represent a distinct biomarker of myocardial cellular turnover, largely independent of conventional biomarkers of injury or functional impairment. This highlights cfDNA as a promising noninvasive tool for early detection and monitoring of subclinical disease processes in HCM. By capturing aspects of myocardial biology that escape standard imaging and laboratory assessment, cfDNA profiling has the potential to refine risk stratification, guide personalized management, and inform mechanistic studies aimed at understanding the cellular drivers of hypertrophy and fibrosis. Future longitudinal and multi-omic studies will be critical to validate these findings and translate cfDNA into clinically actionable insights.

## Methods

### Study design and participants

This was a prospective, single-center study conducted at Oregon Health & Science University (OHSU). All HCM patients were clinically stable and presented for routine follow-up; none had acute illness at the time of enrollment. The few repeated samples from the same HCM patients (3 of the enrolled 54 contributing 2 samples each) were taken at least 1 year apart. Patients with acute myocardial infarction (AMI) were enrolled within 48 hours of percutaneous coronary intervention (PCI). Peripheral venous blood samples were collected in ethylenediaminetetraacetic acid (EDTA) tubes and plasma was isolated by centrifugation at 1,500 rpm for 15 minutes at 4 °C and stored at −80 °C.

### Ethics statement

The study was approved by the OHSU Institutional Review Board. All participants provided written informed consent before enrollment. Participant sex was recorded from the medical record; because of the sample size, analyses were not disaggregated by sex or gender, and sex is reported as a descriptive characteristic (Table 1). Participants did not receive compensation.

### Clinical biomarker analysis

High-sensitivity cardiac troponin I (Beckman cellular assay) and NT-proBNP (Siemens Healthcare) were measured during the same index sample collection.

### Total cfDNA isolation and purification

cfDNA was isolated using a cfDNA purification kit (Active Motif, 25504). Briefly, samples were thawed on ice, and 1.0 mL of plasma was mixed with 1.25 mL of lysis buffer and 25 µL of magnetic beads to lyse residual cellular components and selectively bind cfDNA, respectively.

Bead-bound samples then underwent two rounds of washing using ethanol wash buffer to remove contaminants. Finally, the cfDNA was released from the magnetic beads and eluted in molecular biology-grade water. Yield was assessed using the Qubit 1x dsDNA high sensitivity assay (Fisher Scientific). Extracted and purified cfDNA samples were then stored at −20 °C until library preparation.

### Whole-genome bisulfite sequencing library preparation and sequencing

Whole genome bisulfite sequencing (WGBS) libraries were prepared using the Zymo-Seq Trio WGBS Library Kit (Zymo Research, D5462). For each sample, 5–10 ng of cfDNA was used as input into the library reaction. Bisulfite conversion was carried out in a thermal cycler with lightning conversion reagent (Zymo Research), followed by column-based clean-up and desulfonation. Purified, bisulfite converted DNA then underwent adapter ligation and subsequent purification using Zymo’s Select-a-Size MagBeads. Finally, libraries were polymerase chain reaction (PCR)-amplified for 10 cycles with Zymo’s unique dual index (UDI) primers, and PCR artifacts were removed using the MagBeads (Zymo Research). Final libraries were eluted in DNA elution buffer, quantified, and sequenced 150 bp paired-end on an Illumina NovaSeq platform to generate an average of 274 million raw reads per library.

### Cell-free DNA methylation analysis and deconvolution

Sequencing quality was assessed with FastQC v0.11.9. Raw reads were trimmed with TrimGalore v0.6.7, and trimmed reads were aligned to the human reference genome hg38 with Bismark v0.24.1 using default settings. Duplicates were removed with the deduplicate_bismark script^21–23^.

Cell-type deconvolution of the cfDNA methylome was performed using the UXM fragment-level deconvolution algorithm, which applies non-negative least squares (NNLS) modeling to estimate the relative contributions of 39 primary human cell types based on a reference methylation atlas created by Loyfer et al. This method reconstructs cfDNA methylation patterns as a linear combination of cell-type-specific methylation signatures and has been shown to sensitively identify the tissue of origin using the top 25 unmethylated regions per cell type (a total of 1,246 markers), each comprising multiple CpG sites and selected for their specificity across tissues^18^. For this study, we performed deconvolution using 14 cell types: granulocytes, B cells, monocytes/macrophages, natural killer cells, T cells, endothelial cells, cardiomyocytes, cardiac fibroblasts, kidney epithelium, hepatocytes, alveolar epithelium, bronchial epithelium, skeletal muscle, and smooth muscle.

For the methylation atlas and UXM deconvolution algorithm, the deduplicated BAM files were converted to PAT files using the wgbstools bam2pat function, followed by running UXM deconvolution on the PAT files.

### Validation using independent tissue datasets

WGBS datasets from purified human tissues were obtained from previously published studies, including cardiomyocyte nuclei isolated from left ventricular biopsies^24^, whole blood samples from participants in the London Life Sciences Prospective Population Study (LOLIPOP)^25^, and healthy control liver biopsies obtained during bariatric surgery^26^. Raw sequencing reads were processed using the same pipeline described above.

Liver reference sample L1 was excluded from validation because it contained fewer than 20 million fragments. To normalize sequencing depth, each retained sample was subsampled to 20 million fragments. In silico mixtures were generated by randomly sampling fragments from each tissue dataset in defined proportions (heart:liver:blood ratios of 10:10:80, 5:10:85, 5:5:90) while maintaining equal total depth. Deconvolution was performed in accordance with the previously described approach and predicted tissue fractions were compared with the known mixture proportions.

### Cross-method comparison

To assess the method dependence of the cell type estimates, we compared UXM’s cell-type deconvolution results against an additional reference-based methylation-haplotype-aware algorithm, CelFiE-ISH^27^. The CelFiE-ISH reference atlas was constructed using the same 14 cell types and top 25 atlas markers used for UXM (Supplementary Table 3). The atlas construction workflow was adopted from Unterman et al.^27,28^. Deconvolution was run with the CelFiE-ISH model at default settings. Ground-truth fractions were unavailable for our 50 clinical HCM samples; however, cross-method agreement on the cohort was assessed using Lin’s concordance correlation coefficient (CCC), Pearson r, and Bland–Altman analysis.

### Statistics and reproducibility

Continuous variables are presented as mean ± standard deviation or median with interquartile range, as appropriate, and categorical variables as counts and percentages. Comparisons between groups were performed using Student’s *t*-tests or Wilcoxon rank-sum tests for continuous variables and χ² or Fisher’s exact tests for categorical variables. All tests were two-sided.

Because cfDNA tissue fractions represent compositional data (i.e., proportions constrained to sum to one), fraction values were transformed using a center-log-ratio (CLR) transformation prior to statistical analysis to mitigate compositional bias. Correlations between cfDNA measurements and clinical variables were evaluated using Spearman correlation coefficients. Subsequently, participants were stratified into tertiles based on cfDNA yield or tissue-specific fractions. Clinically relevant variables were then compared across tertiles. Associations between cfDNA measurements and clinical variables were further explored using regression-based modeling approaches. Elastic-net regularized regression models were constructed to evaluate whether combinations of clinical variables could predict cfDNA yield or cardiomyocyte-derived cfDNA fractions. Model performance was evaluated using both in-sample and out-of-sample predictive accuracy. All statistical analyses were performed using Stata version 18 (StataCorp LLC, College Station, TX). *P* values are reported to three decimal places and values <0.005 are shown in bold font in tables. No statistical method was used to predetermine sample size; this was a feasibility study and all samples passing quality control were analyzed. No data were excluded from the analyses. The experiments were not randomized and the investigators were not blinded to allocation during experiments and outcome assessment. Each plasma sample was sequenced once; 57 HCM samples (from 54 patients) and 10 AMI samples were analyzed as biological replicates.

### Reporting summary

Further information on research design is available in the Nature Portfolio Reporting Summary linked to this article.

## Data availability

The raw WGBS sequencing data generated in this study contain identifiable human genetic information and can be requested from the corresponding author, contingent on ethics board approval. The processed per-sample cell-type fraction estimates (UXM and CelFiE-ISH) and de-identified clinical variables used in all analyses are provided in the Source Data file. The publicly available reference WGBS datasets used for validation are available through the NCBI Gene Expression Omnibus under accession numbers GSE186458 (methylation atlas^18^), GSE128734 (LOLIPOP whole blood^25^), and GSE117593 (liver^26^). The cardiomyocyte nuclei WGBS data are available through the NCBI Sequence Read Archive under BioProject accession PRJNA353755^24^. Source data are provided with this paper. Additional information is available from the corresponding authors upon request.

## Acknowledgements

The authors sincerely thank Dr. Mariam Okhovat for her insightful discussions and assistance with study design. We gratefully acknowledge the Epigenetics Consortium at OHSU for executing the computational workflows. Additionally, we thank the ACC server facility at OHSU for generously providing computational resources.

## Author contributions

A.M. and L.C. conceived and designed the study. D.K., B.D., C.E.L., S.A., K.W. and I.J. collected samples and clinical data and performed experiments. B.D. and L.C. performed sequencing data processing and deconvolution analyses. J.W. performed the statistical analyses. D.K., A.M. and L.C. wrote the manuscript. All authors reviewed, edited and approved the final manuscript.

## Competing interests

A.M. reports research grants from Attralus, Bayer, Cytokinetics and Janssen, and personal consulting fees from Cytokinetics, BridgeBio, Pfizer, Ionis, Lexicon, Attralus, Alnylam, Haya, Alexion, Akros, Edgewise, Rocket, Lexeo, Prothena, Bayer, Imbria, Braveheart, BioMarin, AstraZeneca, Avidity, and Neurimmune. The remaining authors declare no competing interests.

## Notes

### Author Declarations

Oregon Health & Science University Institutional Review Board approved the study.

### Summary of Updates

Updated the figures and text. No substantial new data.

### Funding

- NHLBI: 5T32HL166128-02

