## Supplemental Appendix for "Cardiomyocyte-derived cell-free DNA in hypertrophic cardiomyopathy"

**Contents**

Supplementary Fig. 1 | Expected versus predicted tissue fractions in in-silico heart–liver–blood mixtures

Supplementary Fig. 2 | Deconvolution of purified blood, heart and liver reference samples

Supplementary Fig. 3 | Correlation between UXM and CelFiE-ISH estimates across 14 cell types

Supplementary Fig. 4 | Bland–Altman comparison of CelFiE-ISH and UXM estimates across 14 cell types

Supplementary Table 1 | Characteristics of acute myocardial infarction patients

Supplementary Table 2 | Inter-method agreement statistics (UXM versus CelFiE-ISH)

Supplementary Table 3 | WGBS reference samples used to construct the CelFiE-ISH atlas

Supplementary Table 4 | HCM Patient Characteristics by Tertile of Total Heart cfDNA Fraction

Supplementary Table 5 | HCM Patient Characteristics by Tertile of Cardiomyocyte cfDNA Fraction

Supplementary Table 6 | **HCM Characteristics by Tertile of Cardiac Fibroblast-Derived cfDNA Fraction**

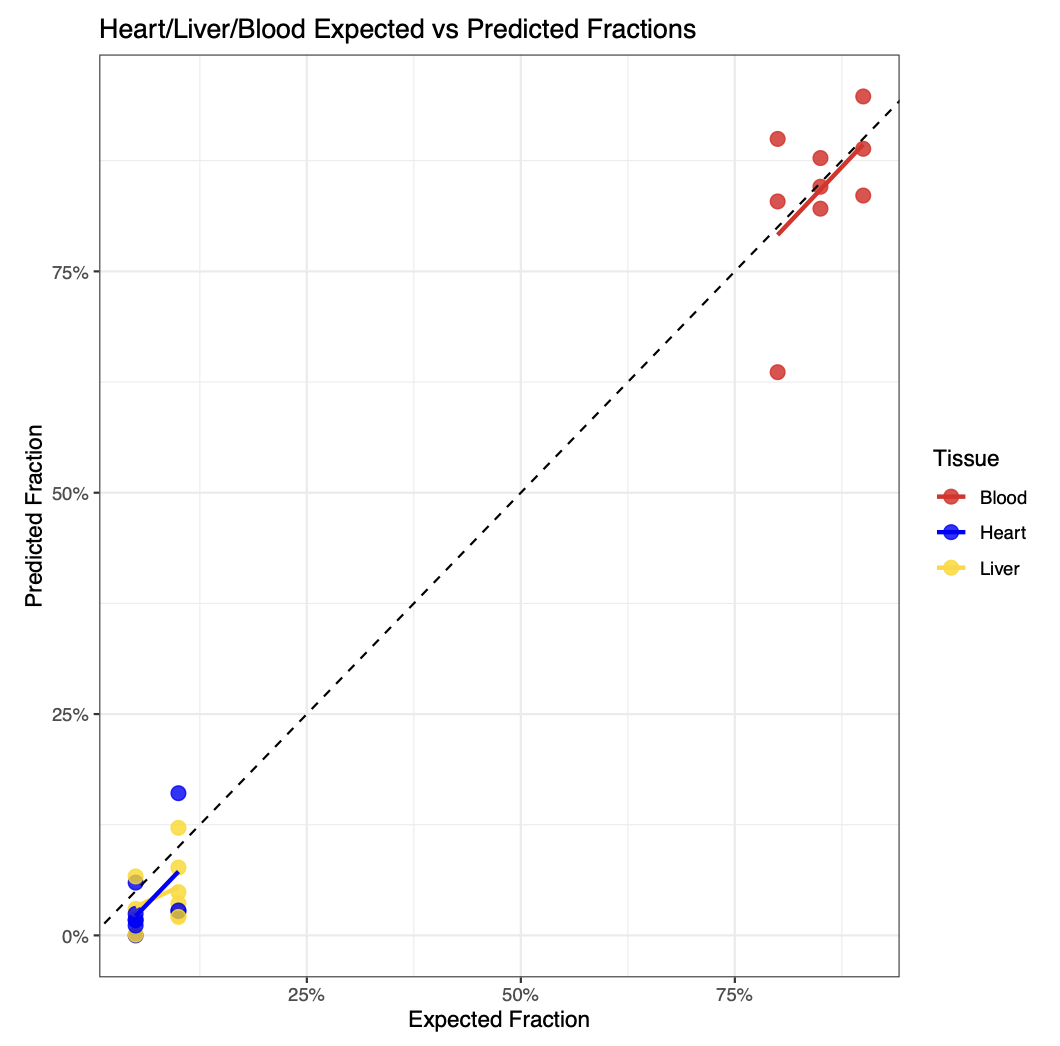

**Supplementary Fig. 1 | Expected versus predicted tissue fractions in in-silico heart–liver–blood mixtures.** WGBS fragments from three heart–liver–blood reference-sample combinations (H1–L2–B1, H2–L3–B2, and H3–L2–B3) were mixed at each of three heart:liver:blood ratios (10:10:80, 5:10:85, and 5:5:90), yielding nine in-silico mixtures. Predicted blood (red), heart (blue), and liver (yellow) fractions are plotted against expected fractions. Colored lines show fitted linear relationships; the dashed line indicates perfect agreement.

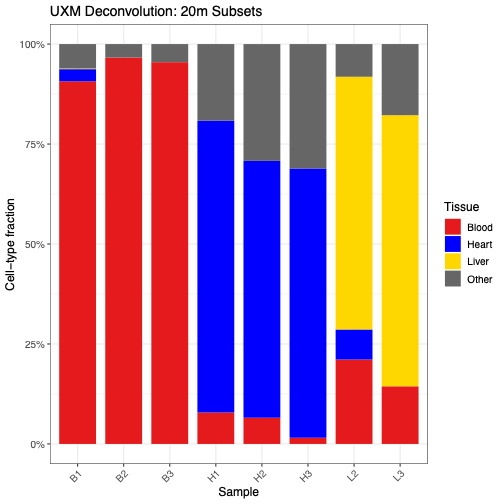

**Supplementary Fig. 2 | Deconvolution of purified blood, heart and liver reference samples with the methylation atlas.** Stacked bars show estimated blood, heart, liver and other tissue fractions in individual blood (B1–B3), heart (H1–H3) and liver (L2–L3) reference samples. "Other" represents contributions assigned to cell types outside the blood, heart and liver categories. L1 did not reach 20 million fragments and was excluded.

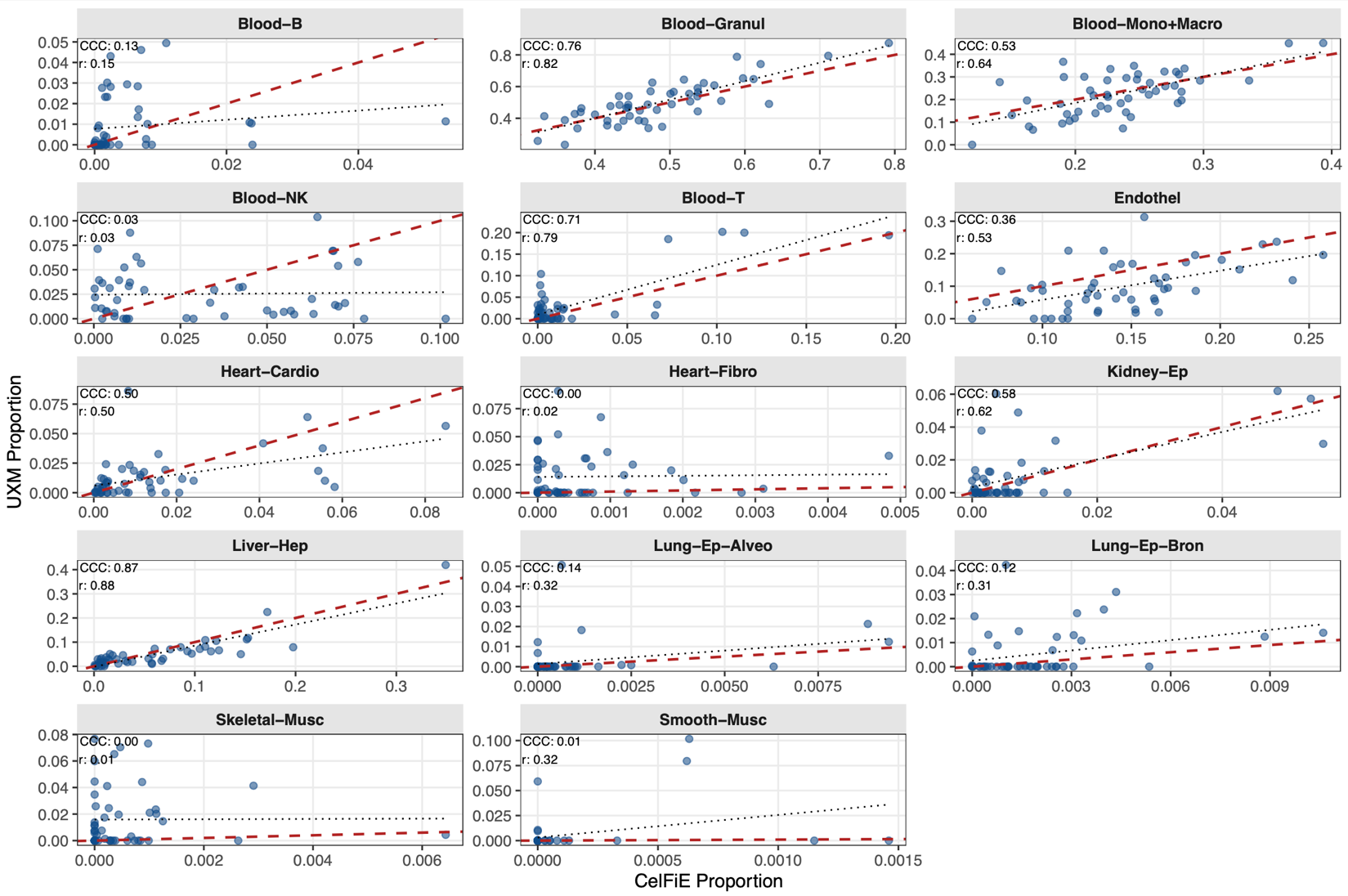

**Supplementary Fig. 3 | Correlation between UXM and CelFiE-ISH cell-type fraction estimates across 14 cell types.** Scatterplots show paired cell-type fraction estimates from CelFiE-ISH (horizontal axis) and UXM (vertical axis) for 50 HCM samples. Each point represents one sample. Red dashed lines indicate perfect agreement between methods, and black dotted lines show fitted linear relationships. Lin's concordance correlation coefficient (CCC) and Pearson's correlation coefficient (r) are reported in each panel.

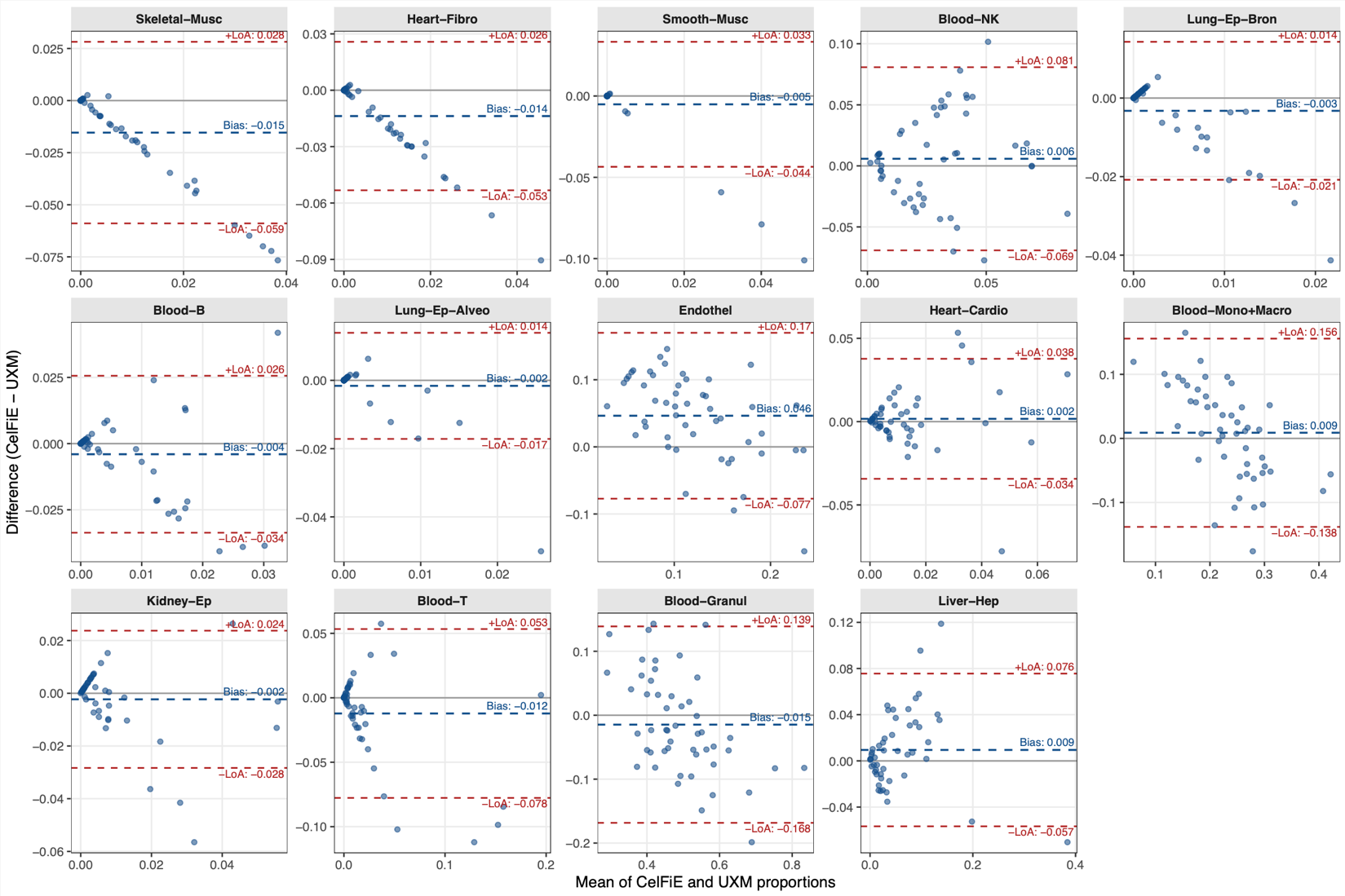

**Supplementary Fig. 4 | Bland–Altman comparison of CelFiE-ISH and UXM estimates across 14 cell types.** Bland–Altman plots compare cell-type fraction estimates across 50 HCM samples. Each point represents one sample, with the mean of the two estimates on the horizontal axis and their difference (CelFiE-ISH − UXM) on the vertical axis. Blue dashed lines indicate mean bias, and red dashed lines indicate the 95% limits of agreement. Gray horizontal lines mark zero difference between methods.

**Supplementary Table 1 | Characteristics of acute myocardial infarction patients**

| ***N (%)***  ***mean ± SD***  ***median (Q1, Q3) [min, max]*** | ***NSTEMI (n=5)*** | ***STEMI (n=5)*** |
| --- | --- | --- |
| Male sex | 3 (60.0%) | 5 (100.0%) |
| Age at sample collection, years | 73.8 ± 4.7 | 67.8 ± 7.7 |
| BMI, kg/m² | 28.5 ± 5.8 | 33.1 ± 5.8 |
| Obese (BMI ≥30 kg/m²) | 3 (60.0%) | 4 (80.0%) |
| Diabetes mellitus | 4 (80.0%) | 3 (60.0%) |
| Obstructive sleep apnea | 0 (0.0%) | 1 (20.0%) |
| Hypertension | 3 (60.0%) | 4 (80.0%) |
| Hemoglobin, g/dL (sex-adjusted) | 13.6 ± 1.3 | 12.9 ± 2.3 |
| Anemic (hemoglobin <12 [F], <13.5 [M] g/dL) | 1 (20.0%) | 2 (40.0%) |
| Creatinine, mg/dL (sex-adjusted) | 1.0 ± 0.2 | 1.1 ± 0.5 |
| Elevated creatinine (>1.0 [F], >1.2 [M] mg/dL) | 1 (20.0%) | 1 (20.0%) |
| LVEF, % | 47.0 ± 12.5 | 41.0 ± 9.6 |
| Atrial fibrillation | 2 (40.0%) | 1 (20.0%) |
| Collection time after symptom onset, hours | 21.7 ± 11.0 | 23.2 ± 9.5 |
| NT-proBNP, pg/mL | 1,570 (765, 1,959) [765, 1,959] | 350 (44, 5,361) [44, 5,361] |
| Troponin at collection, ng/L | 5,926 (5,388, 10,722) [3,332, 15,257] | 49,200 (31,144, 71,993) [23,763, 74,762] |
| Peak troponin, ng/L | 7,514 (6,659, 15,257) [5,775, 25,363] | 99,791 (49,200, 250,795) [35,985, 270,000] |

Clinical, demographic and laboratory characteristics of NSTEMI (*n* = 5) and STEMI (*n* = 5) patients included as positive controls. Data are *n* (%), mean ± SD or median (Q1, Q3) [min, max]. BMI, body mass index; LVEF, left ventricular ejection fraction; NT-proBNP, N-terminal pro-B-type natriuretic peptide.

**Supplementary Table 2 | Inter-method agreement statistics between UXM and CelFiE-ISH**

| **Cell type** | **CCC** | **Cb** | **Pearson r** | **Pearson P** | **Bias** | **Bias 95% CI (lower)** | **Bias 95% CI (upper)** | **LoA (lower)** | **LoA (upper)** |
| --- | --- | --- | --- | --- | --- | --- | --- | --- | --- |
| Liver-Hep | 0.867 | 0.990 | 0.875 | 9.4 × 10⁻17 | 0.009 | -1.1 × 10⁻4 | 0.019 | -0.057 | 0.076 |
| Blood-Granul | 0.761 | 0.932 | 0.817 | 4.6 × 10⁻13 | -0.015 | -0.037 | 0.008 | -0.168 | 0.139 |
| Blood-T | 0.710 | 0.899 | 0.789 | 9.6 × 10⁻12 | -0.012 | -0.022 | -0.003 | -0.078 | 0.053 |
| Kidney-Ep | 0.584 | 0.943 | 0.620 | 1.6 × 10⁻6 | -0.002 | -0.006 | 0.001 | -0.028 | 0.024 |
| Blood-Mono+Macro | 0.535 | 0.837 | 0.639 | 5.9 × 10⁻7 | 0.009 | -0.012 | 0.030 | -0.138 | 0.156 |
| Heart-Cardio | 0.501 | 0.993 | 0.505 | 1.8 × 10⁻4 | 0.002 | -0.004 | 0.007 | -0.034 | 0.038 |
| Endothel | 0.362 | 0.679 | 0.533 | 6.7 × 10⁻5 | 0.046 | 0.028 | 0.064 | -0.077 | 0.170 |
| Lung-Ep-Alveo | 0.138 | 0.434 | 0.317 | 0.025 | -0.002 | -0.004 | 6.4 × 10⁻4 | -0.017 | 0.014 |
| Blood-B | 0.131 | 0.872 | 0.150 | 0.298 | -0.004 | -0.008 | 2.9 × 10⁻4 | -0.034 | 0.026 |
| Lung-Ep-Bron | 0.117 | 0.375 | 0.313 | 0.027 | -0.003 | -0.006 | -7.0 × 10⁻4 | -0.021 | 0.014 |
| Blood-NK | 0.029 | 0.972 | 0.030 | 0.836 | 0.006 | -0.005 | 0.017 | -0.069 | 0.081 |
| Smooth-Musc | 0.009 | 0.027 | 0.325 | 0.021 | -0.005 | -0.011 | 4.6 × 10⁻4 | -0.044 | 0.033 |
| Heart-Fibro | 0.002 | 0.063 | 0.024 | 0.870 | -0.014 | -0.019 | -0.008 | -0.053 | 0.026 |
| Skeletal-Musc | 3.5 × 10⁻4 | 0.063 | 0.006 | 0.970 | -0.015 | -0.022 | -0.009 | -0.059 | 0.028 |

Agreement between UXM and CelFiE-ISH cell-type fraction estimates across 50 HCM samples, assessed using Lin's concordance correlation coefficient (CCC), the bias-correction factor (Cb), Pearson correlation, mean bias (CelFiE-ISH − UXM) with 95% confidence interval, and Bland–Altman 95% limits of agreement (LoA). Values rounded to three decimal places; values below 0.001 are given in scientific notation.

**Supplementary Table 3 | WGBS reference samples used to construct the CelFiE-ISH atlas**

| **Name** | **Group** |
| --- | --- |
| GSE186458/GSM5652317_Blood-B-Z000000UB.hg38.pat.gz | Blood-B |
| GSE186458/GSM5652318_Blood-B-Z000000UR.hg38.pat.gz | Blood-B |
| GSE186458/GSM5652319_Blood-B-Mem-Z0000041J.hg38.pat.gz | Blood-B |
| GSE186458/GSM5652320_Blood-B-Mem-Z0000041K.hg38.pat.gz | Blood-B |
| GSE186458/GSM5652314_Blood-Granulocytes-Z000000UD.hg38.pat.gz | Blood-Granul |
| GSE186458/GSM5652315_Blood-Granulocytes-Z000000UT.hg38.pat.gz | Blood-Granul |
| GSE186458/GSM5652303_Blood-Monocytes-Z000000U3.hg38.pat.gz | Blood-Mono+Macro |
| GSE186458/GSM5652304_Blood-Monocytes-Z000000UH.hg38.pat.gz | Blood-Mono+Macro |
| GSE186458/GSM5652305_Colon-Macrophages-Z00000444.hg38.pat.gz | Blood-Mono+Macro |
| GSE186458/GSM5652306_Colon-Macrophages-Z00000446.hg38.pat.gz | Blood-Mono+Macro |
| GSE186458/GSM5652307_Liver-Macrophages-Z0000043P.hg38.pat.gz | Blood-Mono+Macro |
| GSE186458/GSM5652308_Lung-Alveolar-Macrophages-Z00000448.hg38.pat.gz | Blood-Mono+Macro |
| GSE186458/GSM5652309_Lung-Alveolar-Macrophages-Z0000044C.hg38.pat.gz | Blood-Mono+Macro |
| GSE186458/GSM5652310_Lung-Interstitial-Macrophages-Z00000447.hg38.pat.gz | Blood-Mono+Macro |
| GSE186458/GSM5652311_Lung-Interstitial-Macrophages-Z0000044D.hg38.pat.gz | Blood-Mono+Macro |
| GSE186458/GSM5652312_Lung-Interstitial-Macrophages-Z0000044E.hg38.pat.gz | Blood-Mono+Macro |
| GSE186458/GSM5652300_Blood-NK-Z000000U1.hg38.pat.gz | Blood-NK |
| GSE186458/GSM5652301_Blood-NK-Z000000UF.hg38.pat.gz | Blood-NK |
| GSE186458/GSM5652278_Blood-T-CD3-Z000000UP.hg38.pat.gz | Blood-T |
| GSE186458/GSM5652279_Blood-T-CD4-Z000000TT.hg38.pat.gz | Blood-T |
| GSE186458/GSM5652280_Blood-T-CD4-Z000000U7.hg38.pat.gz | Blood-T |
| GSE186458/GSM5652281_Blood-T-CD4-Z000000UM.hg38.pat.gz | Blood-T |
| GSE186458/GSM5652282_Blood-T-CD8-Z000000TR.hg38.pat.gz | Blood-T |
| GSE186458/GSM5652283_Blood-T-CD8-Z000000U5.hg38.pat.gz | Blood-T |
| GSE186458/GSM5652284_Blood-T-CD8-Z000000UK.hg38.pat.gz | Blood-T |
| GSE186458/GSM5652285_Blood-T-CenMem-CD4-Z00000417.hg38.pat.gz | Blood-T |
| GSE186458/GSM5652286_Blood-T-CenMem-CD4-Z0000041D.hg38.pat.gz | Blood-T |
| GSE186458/GSM5652287_Blood-T-CenMem-CD4-Z0000041N.hg38.pat.gz | Blood-T |
| GSE186458/GSM5652288_Blood-T-Eff-CD8-Z00000419.hg38.pat.gz | Blood-T |
| GSE186458/GSM5652289_Blood-T-Eff-CD8-Z0000041F.hg38.pat.gz | Blood-T |
| GSE186458/GSM5652290_Blood-T-Eff-CD8-Z0000041Q.hg38.pat.gz | Blood-T |
| GSE186458/GSM5652291_Blood-T-EffMem-CD4-Z00000416.hg38.pat.gz | Blood-T |
| GSE186458/GSM5652292_Blood-T-EffMem-CD4-Z0000041C.hg38.pat.gz | Blood-T |
| GSE186458/GSM5652293_Blood-T-EffMem-CD4-Z0000041M.hg38.pat.gz | Blood-T |
| GSE186458/GSM5652294_Blood-T-EffMem-CD8-Z0000041A.hg38.pat.gz | Blood-T |
| GSE186458/GSM5652295_Blood-T-EffMem-CD8-Z0000041G.hg38.pat.gz | Blood-T |
| GSE186458/GSM5652296_Blood-T-Naive-CD4-Z0000041E.hg38.pat.gz | Blood-T |
| GSE186458/GSM5652297_Blood-T-Naive-CD8-Z0000041B.hg38.pat.gz | Blood-T |
| GSE186458/GSM5652298_Blood-T-Naive-CD8-Z0000041H.hg38.pat.gz | Blood-T |
| GSE186458/GSM5652180_Aorta-Endothel-Z0000043G.hg38.pat.gz | Endothel |
| GSE186458/GSM5652184_Kidney-Glomerular-Endothel-Z000000Q5.hg38.pat.gz | Endothel |
| GSE186458/GSM5652185_Kidney-Glomerular-Endothel-Z00000443.hg38.pat.gz | Endothel |
| GSE186458/GSM5652186_Kidney-Glomerular-Endothel-Z0000045J.hg38.pat.gz | Endothel |
| GSE186458/GSM5652187_Kidney-Tubular-Endothel-Z000000PX.hg38.pat.gz | Endothel |
| GSE186458/GSM5652188_Kidney-Tubular-Endothel-Z000000Q3.hg38.pat.gz | Endothel |
| GSE186458/GSM5652189_Kidney-Tubular-Endothel-Z0000042R.hg38.pat.gz | Endothel |
| GSE186458/GSM5652190_Liver-Endothelium-Z000000RB.hg38.pat.gz | Endothel |
| GSE186458/GSM5652191_Lung-Alveolar-Endothel-Z000000Q1.hg38.pat.gz | Endothel |
| GSE186458/GSM5652192_Lung-Alveolar-Endothel-Z000000QK.hg38.pat.gz | Endothel |
| GSE186458/GSM5652193_Lung-Alveolar-Endothel-Z0000045H.hg38.pat.gz | Endothel |
| GSE186458/GSM5652194_Pancreas-Endothel-Z0000042D.hg38.pat.gz | Endothel |
| GSE186458/GSM5652195_Pancreas-Endothel-Z0000042X.hg38.pat.gz | Endothel |
| GSE186458/GSM5652196_Pancreas-Endothel-Z00000430.hg38.pat.gz | Endothel |
| GSE186458/GSM5652197_Pancreas-Islet-Endothel-Z0000042Y.hg38.pat.gz | Endothel |
| GSE186458/GSM5652181_Saphenous-Vein-Endothel-Z000000RM.hg38.pat.gz | Endothel |
| GSE186458/GSM5652182_Saphenous-Vein-Endothel-Z000000S7.hg38.pat.gz | Endothel |
| GSE186458/GSM5652183_Saphenous-Vein-Endothel-Z000000SB.hg38.pat.gz | Endothel |
| GSE186458/GSM5652213_Heart-Cardiomyocyte-Z0000044K.hg38.pat.gz | Heart-Cardio |
| GSE186458/GSM5652214_Heart-Cardiomyocyte-Z0000044N.hg38.pat.gz | Heart-Cardio |
| GSE186458/GSM5652215_Heart-Cardiomyocyte-Z0000044P.hg38.pat.gz | Heart-Cardio |
| GSE186458/GSM5652216_Heart-Cardiomyocyte-Z0000044Q.hg38.pat.gz | Heart-Cardio |
| GSE186458/GSM5652217_Heart-Cardiomyocyte-Z0000044R.hg38.pat.gz | Heart-Cardio |
| GSE186458/GSM5652201_Heart-Fibroblasts-Z0000041V.hg38.pat.gz | Heart-Fibro |
| GSE186458/GSM5652202_Heart-Fibroblasts-Z0000041W.hg38.pat.gz | Heart-Fibro |
| GSE186458/GSM5652203_Heart-Fibroblasts-Z0000041X.hg38.pat.gz | Heart-Fibro |
| GSE186458/GSM5652257_Kidney-Glomerular-Epithelial-Z0000045L.hg38.pat.gz | Kidney-Ep |
| GSE186458/GSM5652261_Kidney-Glomerular-Podocytes-Z0000042W.hg38.pat.gz | Kidney-Ep |
| GSE186458/GSM5652262_Kidney-Glomerular-Podocytes-Z00000441.hg38.pat.gz | Kidney-Ep |
| GSE186458/GSM5652263_Kidney-Glomerular-Podocytes-Z00000442.hg38.pat.gz | Kidney-Ep |
| GSE186458/GSM5652258_Kidney-Tubular-Epithelial-Z000000QH.hg38.pat.gz | Kidney-Ep |
| GSE186458/GSM5652259_Kidney-Tubular-Epithelial-Z0000043Z.hg38.pat.gz | Kidney-Ep |
| GSE186458/GSM5652260_Kidney-Tubular-Epithelial-Z00000440.hg38.pat.gz | Kidney-Ep |
| GSE186458/GSM5652234_Liver-Hepatocytes-Z000000T3.hg38.pat.gz | Liver-Hep |
| GSE186458/GSM5652235_Liver-Hepatocytes-Z0000043Q.hg38.pat.gz | Liver-Hep |
| GSE186458/GSM5652236_Liver-Hepatocytes-Z0000044H.hg38.pat.gz | Liver-Hep |
| GSE186458/GSM5652237_Liver-Hepatocytes-Z0000044M.hg38.pat.gz | Liver-Hep |
| GSE186458/GSM5652238_Liver-Hepatocytes-Z00000431.hg38.pat.gz | Liver-Hep |
| GSE186458/GSM5652206_Skeletal-Muscle-Z00000429.hg38.pat.gz | Skeletal-Musc |
| GSE186458/GSM5652209_Bladder-Smooth-Muscle-Z0000041Z.hg38.pat.gz | Smooth-Musc |
| GSE186458/GSM5652208_Coronary-Artery-Smooth-Muscle-Z00000420.hg38.pat.gz | Smooth-Musc |
| GSE186458/GSM5652211_Lung-Bronchus-Smooth-Muscle-Z00000421.hg38.pat.gz | Smooth-Musc |
| GSE186458/GSM5652210_Prostate-Smooth-Muscle-Z0000041Y.hg38.pat.gz | Smooth-Musc |
| GSE186458/GSM5652355_Lung-Alveolar-Epithelial-Z000000VC.hg38.pat.gz | Lung-Ep-Alveo |
| GSE186458/GSM5652356_Lung-Alveolar-Epithelial-Z000000VE.hg38.pat.gz | Lung-Ep-Alveo |
| GSE186458/GSM5652336_Lung-Bronchus-Epithelial-Z000000RZ.hg38.pat.gz | Lung-Ep-Bron |
| GSE186458/GSM5652337_Lung-Bronchus-Epithelial-Z000000S5.hg38.pat.gz | Lung-Ep-Bron |

Reference WGBS sample identifiers (GEO series GSE186458^18^) and corresponding cell-type group assignments used to construct the CelFiE-ISH reference atlas.

**Supplementary Table 4 | HCM Patient Characteristics by Tertile of Total Heart cfDNA Fraction**

| *N (%)*  *mean ± SD*  *median (Q1, Q3) [min, max]* | **First Tertile**  *n*=19 | **Second Tertile**  *n*=19 | **Third Tertile**  *n*=19 | *p-value* |
| --- | --- | --- | --- | --- |
| Male sex | 14 (73.7%) | 13 (68.4%) | 10 (52.6%) | 0.472 |
| Age at sample collection | 50.5 ± 19.5 | 50.3 ± 12.8 | 38.3 ± 14.4 | 0.042 |
| Age > 50 years old | 10 (52.6%) | 9 (47.4%) | 4 (21.1%) | 0.119 |
| BMI (kg/m^2^) | 28.4 ± 4.6 | 32.3 ± 4.9 | 34.0 ± 5.6 | **0.004** |
| Obese (BMI ≥ 30) | 5 (26.3%) | 14 (73.7%) | 14 (73.7%) | **0.004** |
| Diabetes mellitus | 1 (5.3%) | 0 (0.0%) | 1 (5.3%) | >0.999 |
| Coronary artery disease | 2 (10.5%) | 3 (15.8%) | 0 (0.0%) | 0.351 |
| Obstructive sleep apnea | 0 (0.0%) | 5 (26.3%) | 3 (15.8%) | 0.079 |
| Hypertension | 5 (26.3%) | 8 (42.1%) | 2 (10.5%) | 0.101 |
| Years since HCM diagnosis | 5 (1, 10)  [0, 20] | 5 (3, 8)  [0, 34] | 10 (1, 21)  [0, 26] | 0.334 |
| NYHA class |  |  |  | >0.999 |
| *I* | 4 (21.1%) | 5 (26.3%) | 4 (21.1%) |  |
| *II* | 11 (57.9%) | 10 (52.6%) | 10 (52.6%) |  |
| *III/IV* | 4 (21.1%) | 4 (21.1%) | 5 (26.3%) |  |
| Family history of HCM | 9 (47.4%) | 9 (47.4%) | 9 (47.4%) | >0.999 |
| Pathological genetic mutation |  |  |  | 0.227 |
| *no* | 5 (26.3%) | 8 (42.1%) | 2 (10.5%) |  |
| *yes* | 9 (47.4%) | 5 (26.3%) | 10 (52.6%) |  |
| *unknown* | 5 (26.3%) | 6 (31.6%) | 7 (36.8%) |  |
| Late gadolinium enhancement |  |  |  | 0.039 |
| *no* | 4 (21.1%) | 6 (31.6%) | 2 (10.5%) |  |
| *yes* | 12 (63.2%) | 13 (68.4%) | 10 (52.6%) |  |
| *unknown* | 3 (15.8%) | 0 (0.0%) | 7 (36.8%) |  |
| Degree of fibrosis |  |  |  | 0.270 |
| *None* | 4 (25.0%) | 6 (31.6%) | 2 (16.7%) |  |
| *Mild* | 8 (50.0%) | 12 (63.2%) | 6 (50.0%) |  |
| *Moderate* | 2 (12.5%) | 1 (5.3%) | 4 (33.3%) |  |
| *Severe* | 2 (12.5%) | 0 (0.0%) | 0 (0.0%) |  |
| Symptomatic | 13 (68.4%) | 15 (78.9%) | 14 (73.7%) | 0.929 |
| Obstruction | 4 (21.1%) | 11 (57.9%) | 11 (57.9%) | 0.031 |
| AFIB | 1 (5.3%) | 1 (5.3%) | 3 (15.8%) | 0.602 |
| Implantable cardioverter-defibrillator | 10 (52.6%) | 6 (31.6%) | 9 (47.4%) | 0.498 |
| Myectomy | 4 (21.1%) | 3 (15.8%) | 6 (31.6%) | 0.625 |
| Hemoglobin (g/dL) (sex-adjusted) | 14.1 ± 1.1 | 14.8 ± 0.6 | 14.8 ± 1.2 | 0.066 |
| Anemic (hemoglobin < 12[F],13.5[M]) | 3 (15.8%) | 0 (0.0%) | 2 (10.5%) | 0.351 |
| Creatinine (mg/dL) (sex-adjusted) | 1.0 ± 0.1 | 0.9 ± 0.2 | 0.8 ± 0.1 | 0.056 |
| Elevated creatinine (>1.0[F],1.2[M]) | 4 (21.1%) | 2 (10.5%) | 1 (5.3%) | 0.476 |
| NT-proBNP (pg/mL) | 668 (125, 1229)  [44, 4351] | 334 (106, 501)  [29, 1275] | 308 (213, 1059)  [27, 3014] | 0.126 |
| Elevated NT-proBNP (>300) | 14 (73.7%) | 10 (52.6%) | 11 (57.9%) | 0.485 |
| Troponin at collection (ng/L) | 54 (9, 130)  [3, 3361] | 8 (7, 57)  [4, 676] | 20 (7, 172)  [3, 7775] | 0.485 |
| Elevated troponin (>10) | 12 (63.2%) | 9 (47.4%) | 11 (57.9%) | 0.711 |

Clinical, demographic, imaging, and laboratory characteristics of HCM samples stratified by total heart-derived cfDNA fraction tertile (*n* = 19 per tertile). Data are *n* (%), mean ± SD or median (Q1, Q3) [min, max]. BMI, body mass index; LVEF, left ventricular ejection fraction; NT-proBNP, N-terminal pro-B-type natriuretic peptide. Two-sided *P* values; *P* < 0.005 in bold.

**Supplementary Table 5 | HCM Patient Characteristics by Tertile of Cardiomyocyte cfDNA Fraction**

| *N (%)*  *mean ± SD*  *median (Q1, Q3) [min, max]* | **First Tertile**  *n*=19 | **Second Tertile**  *n*=19 | **Third Tertile**  *n*=19 | *p-value* |
| --- | --- | --- | --- | --- |
| Male sex | 12 (63.2%) | 15 (78.9%) | 10 (52.6%) | 0.272 |
| Age at sample collection | 46.7 ± 20.6 | 47.3 ± 15.1 | 45.1 ± 14.2 | 0.946 |
| Age > 50 years old | 8 (42.1%) | 7 (36.8%) | 8 (42.1%) | >0.999 |
| BMI (kg/m^2^) | 28.2 ± 5.0 | 32.2 ± 4.5 | 34.2 ± 5.4 | **0.001** |
| Obese (BMI ≥ 30) | 4 (21.1%) | 14 (73.7%) | 15 (78.9%) | **<0.001** |
| Diabetes mellitus | 1 (5.3%) | 0 (0.0%) | 1 (5.3%) | >0.999 |
| Coronary artery disease | 2 (10.5%) | 2 (10.5%) | 1 (5.3%) | >0.999 |
| Obstructive sleep apnea | 1 (5.3%) | 1 (5.3%) | 6 (31.6%) | 0.038 |
| Hypertension | 4 (21.1%) | 7 (36.8%) | 4 (21.1%) | 0.596 |
| Years since HCM diagnosis | 5 (4, 12)  [0, 25] | 5 (0, 18)  [0, 25] | 8 (1, 17)  [0, 34] | 0.617 |
| NYHA class |  |  |  | 0.231 |
| *I* | 5 (26.3%) | 5 (26.3%) | 3 (15.8%) |  |
| *II* | 12 (63.2%) | 11 (57.9%) | 8 (42.1%) |  |
| *III/IV* | 2 (10.5%) | 3 (15.8%) | 8 (42.1%) |  |
| Family history of HCM | 8 (42.1%) | 11 (57.9%) | 8 (42.1%) | 0.668 |
| Pathological genetic mutation |  |  |  | 0.581 |
| *no* | 6 (31.6%) | 6 (31.6%) | 3 (15.8%) |  |
| *yes* | 9 (47.4%) | 7 (36.8%) | 8 (42.1%) |  |
| *unknown* | 4 (21.1%) | 6 (31.6%) | 8 (42.1%) |  |
| Late gadolinium enhancement |  |  |  | 0.086 |
| *no* | 4 (21.1%) | 5 (26.3%) | 3 (15.8%) |  |
| *yes* | 9 (47.4%) | 14 (73.7%) | 12 (63.2%) |  |
| *unknown* | 6 (31.6%) | 0 (0.0%) | 4 (21.1%) |  |
| Degree of fibrosis |  |  |  | 0.203 |
| *None* | 4 (30.8%) | 5 (26.3%) | 3 (20.0%) |  |
| *Mild* | 7 (53.8%) | 9 (47.4%) | 10 (66.7%) |  |
| *Moderate* | 0 (0.0%) | 5 (26.3%) | 2 (13.3%) |  |
| *Severe* | 2 (15.4%) | 0 (0.0%) | 0 (0.0%) |  |
| Symptomatic | 11 (57.9%) | 15 (78.9%) | 16 (84.2%) | 0.242 |
| Obstruction | 5 (26.3%) | 9 (47.4%) | 12 (63.2%) | 0.088 |
| AFIB | 1 (5.3%) | 0 (0.0%) | 4 (21.1%) | 0.114 |
| Implantable cardioverter-defibrillator | 11 (57.9%) | 8 (42.1%) | 6 (31.6%) | 0.300 |
| Myectomy | 7 (36.8%) | 2 (10.5%) | 4 (21.1%) | 0.185 |
| Hemoglobin (g/dL) (sex-adjusted) | 14.3 ± 1.1 | 14.5 ± 1.0 | 14.9 ± 0.9 | 0.164 |
| Anemic (hemoglobin < 12[F],13.5[M]) | 3 (15.8%) | 1 (5.3%) | 1 (5.3%) | 0.602 |
| Creatinine (mg/dL) (sex-adjusted) | 0.9 ± 0.2 | 0.9 ± 0.2 | 0.8 ± 0.1 | 0.202 |
| Elevated creatinine (>1.0[F],1.2[M]) | 3 (15.8%) | 3 (15.8%) | 1 (5.3%) | 0.678 |
| NT-proBNP (pg/mL) | 428 (180, 755)  [34, 3966] | 501 (79, 898)  [29, 4351] | 304 (135, 871)  [27, 3014] | 0.827 |
| Elevated NT-proBNP (>300) | 13 (68.4%) | 12 (63.2%) | 10 (52.6%) | 0.702 |
| Troponin at collection (ng/L) | 50 (7, 102)  [3, 468] | 25 (8, 676)  [3, 3361] | 8 (5, 139)  [4, 7775] | 0.293 |
| Elevated troponin (>10) | 12 (63.2%) | 13 (68.4%) | 7 (36.8%) | 0.127 |

Clinical, demographic, imaging, and laboratory characteristics of HCM samples stratified by cardiomyocyte-derived cfDNA fraction tertile (*n* = 19 per tertile). Data are *n* (%), mean ± SD or median (Q1, Q3) [min, max]. BMI, body mass index; LVEF, left ventricular ejection fraction; NT-proBNP, N-terminal pro-B-type natriuretic peptide. Two-sided *P* values; *P* < 0.005 in bold.

| *N (%)*  *mean ± SD*  *median (Q1, Q3) [min, max]* | **First Tertile**  *n*=19 | **Second Tertile**  *n*=19 | **Third Tertile**  *n*=19 | *p-value* |
| --- | --- | --- | --- | --- |
| Male sex | 11 (57.9%) | 13 (68.4%) | 13 (68.4%) | 0.829 |
| Age at sample collection | 53.2 ± 16.6 | 43.3 ± 16.5 | 42.6 ± 15.2 | 0.105 |
| Age > 50 years old | 10 (52.6%) | 7 (36.8%) | 6 (31.6%) | 0.491 |
| BMI (kg/m^2^) | 31.2 ± 6.4 | 31.1 ± 5.0 | 32.4 ± 5.1 | 0.641 |
| Obese (BMI ≥ 30) | 10 (52.6%) | 12 (63.2%) | 11 (57.9%) | 0.943 |
| Diabetes mellitus | 1 (5.3%) | 0 (0.0%) | 1 (5.3%) | >0.999 |
| Coronary artery disease | 2 (10.5%) | 1 (5.3%) | 2 (10.5%) | >0.999 |
| Obstructive sleep apnea | 3 (15.8%) | 1 (5.3%) | 4 (21.1%) | 0.503 |
| Hypertension | 5 (26.3%) | 5 (26.3%) | 5 (26.3%) | >0.999 |
| Years since HCM diagnosis | 8 (4, 14)  [0, 34] | 5 (0, 10)  [0, 25] | 5 (1, 21)  [0, 26] | 0.209 |
| NYHA class |  |  |  | 0.924 |
| *I* | 3 (15.8%) | 5 (26.3%) | 5 (26.3%) |  |
| *II* | 12 (63.2%) | 10 (52.6%) | 9 (47.4%) |  |
| *III/IV* | 4 (21.1%) | 4 (21.1%) | 5 (26.3%) |  |
| Family history of HCM | 6 (31.6%) | 12 (63.2%) | 9 (47.4%) | 0.170 |
| Pathological genetic mutation |  |  |  | 0.104 |
| *no* | 5 (26.3%) | 6 (31.6%) | 4 (21.1%) |  |
| *yes* | 4 (21.1%) | 10 (52.6%) | 10 (52.6%) |  |
| *unknown* | 10 (52.6%) | 3 (15.8%) | 5 (26.3%) |  |
| Late gadolinium enhancement |  |  |  | 0.073 |
| *no* | 5 (26.3%) | 4 (21.1%) | 3 (15.8%) |  |
| *yes* | 10 (52.6%) | 15 (78.9%) | 10 (52.6%) |  |
| *unknown* | 4 (21.1%) | 0 (0.0%) | 6 (31.6%) |  |
| Degree of fibrosis |  |  |  | 0.747 |
| *None* | 5 (33.3%) | 4 (21.1%) | 3 (23.1%) |  |
| *Mild* | 9 (60.0%) | 10 (52.6%) | 7 (53.8%) |  |
| *Moderate* | 1 (6.7%) | 3 (15.8%) | 3 (23.1%) |  |
| *Severe* | 0 (0.0%) | 2 (10.5%) | 0 (0.0%) |  |
| Symptomatic | 14 (73.7%) | 15 (78.9%) | 13 (68.4%) | 0.929 |
| Obstruction | 9 (47.4%) | 6 (31.6%) | 11 (57.9%) | 0.303 |
| AFIB | 1 (5.3%) | 1 (5.3%) | 3 (15.8%) | 0.602 |
| Implantable cardioverter-defibrillator | 8 (42.1%) | 9 (47.4%) | 8 (42.1%) | >0.999 |
| Myectomy | 3 (15.8%) | 4 (21.1%) | 6 (31.6%) | 0.625 |
| Hemoglobin (g/dL) (sex-adjusted) | 14.1 ± 1.0 | 14.6 ± 1.0 | 14.9 ± 1.0 | 0.036 |
| Anemic (hemoglobin < 12[F],13.5[M]) | 2 (10.5%) | 2 (10.5%) | 1 (5.3%) | >0.999 |
| Creatinine (mg/dL) (sex-adjusted) | 0.9 ± 0.2 | 0.9 ± 0.1 | 0.8 ± 0.1 | 0.165 |
| Elevated creatinine (>1.0[F],1.2[M]) | 4 (21.1%) | 2 (10.5%) | 1 (5.3%) | 0.476 |
| NT-proBNP (pg/mL) | 457 (213, 871)  [70, 4351] | 501 (135, 898)  [44, 1916] | 308 (44, 746)  [27, 3014] | 0.574 |
| Elevated NT-proBNP (>300) | 13 (68.4%) | 11 (57.9%) | 11 (57.9%) | 0.835 |
| Troponin at collection (ng/L) | 10 (8, 118)  [3, 2822] | 25 (8, 130)  [3, 7775] | 23 (5, 139)  [3, 1092] | 0.681 |
| Elevated troponin (>10) | 8 (42.1%) | 12 (63.2%) | 12 (63.2%) | 0.358 |

**Supplementary Table 6 | HCM Characteristics by Tertile of Cardiac Fibroblast-Derived cfDNA Fraction**

Clinical, demographic, imaging, and laboratory characteristics of HCM samples stratified by cardiac fibroblast-derived cfDNA fraction tertile (*n* = 19 per tertile). Data are *n* (%), mean ± SD or median (Q1, Q3) [min, max]. BMI, body mass index; LVEF, left ventricular ejection fraction; NT-proBNP, N-terminal pro-B-type natriuretic peptide. Two-sided *P* values; *P* < 0.005 in bold.
